# Effect of Data Preprocessing Decisions on Sepsis-3 Labels in MIMIC-III and eICU-CRD: A Sensitivity Analysis

**DOI:** 10.64898/2026.11.12.26367959

**Authors:** Falk Meyer-Eschenbach, Johannes v. Stoephasius, Anatol-Fiete Näher, Fabian Kohlbacher, Severin Kohler, Elias Grünewald, Nicolas Frey, Andreas Edel, Jon Barrenetxea, Felix Balzer, Oliver Kumpf

## Abstract

**Objective:** Published Sepsis-3 detection rates range from 3.4 % to 65.2 % in MIMIC-III and from 9.8 % to 47.9 % in eICU-CRD. We tested whether data preprocessing decisions alone produce differences of this magnitude when data, cohort and sepsis definition are held fixed.

**Materials and Methods:** Using the MIMIC-III and eICU-CRD databases we developed and applied a configurable pipeline (medalabel) that stepwise maps the databases into the OMOP Common Data Model, standardizes and cleans the data, and preprocesses them to compute Sepsis-3 labels. Each setting of the preprocessing represents one decision of published Sepsis-3 implementations, and settings can be freely combined. On fixed cohorts of first ICU stays (13,388 in MIMIC-III, 94,782 in eICU-CRD), we changed one setting of a reference configuration at a time (51 variants in five preprocessing domains) and compared detection rates and septic stays (Jaccard index) with the reference. We approximately reconstructed six published pipelines and combined the settings with the largest effects in pairs to test whether their effects add up. The time a value is measured can differ from the time it is documented and thus available for automated analysis. Beyond the usual retrospective view based on measurement time, the pipeline optionally combines both time points to simulate a real-time system.

**Results:** The reference configuration labelled 31.5 % of ICU stays in MIMIC-III and 8.2 % in eICU-CRD as septic. Changing a single setting resulted in sepsis detection rates of 15.3–58.6 % in MIMIC-III and 4.6– 17.2 % in eICU-CRD. The largest shifts from the reference, all in the same direction in both databases, came from the SOFA baseline, up to +27.1 percentage points (pp) in MIMIC-III and +9.0 pp in eICU-CRD, from disabling imputation (−16.3 and −2.5 pp), the temporal window (+12.6 and +5.3 pp) and aggregation (−10.1 and −0.8 pp); clinician-validated parameter sets had little effect (−4.8 and −1.1 pp). Even variants that changed the rate by less than 2 pp labelled partly different patients, with Jaccard indices as low as 0.67 in MIMIC-III and 0.56 in eICU-CRD (1 = identical septic stays). Reconstructions deviated from the published rates by 2–10 pp. Two settings applied together deviated from the sum of their single effects by 0.9–11.8 pp in MIMIC-III and by up to 4.9 pp in eICU-CRD. Simulating a real-time system lowered the MIMIC-III rate by 5.0 pp and delayed the label in 45 % of the stays that remained septic.

**Discussion:** With the infection criterion held fixed, preprocessing decisions alone can produce a considerable part of the variation between published sepsis detection rates. Similar rates can stem from different patients, which makes studies and prediction models difficult to compare, and retrospective labels rely on values that a real-time system would not yet have.

**Conclusion:** Sepsis-3 labels from health records are only comparable if their data preprocessing is known. Configurable pipelines allow quantifying the effect of these decisions, computing alternative labels on the same data and, in future, validating them against clinician-adjudicated cases.

## Introduction

Sepsis caused an estimated 11 million deaths in 2017, corresponding to 19 % of all deaths worldwide [1, 2], and it causes substantial in-hospital treatment costs [3]. In intensive care units, 29.5 % of patients are affected, with a hospital mortality of 35.3 % [4, 5]. The Sepsis-3 consensus defines sepsis as life-threatening organ dysfunction caused by a dysregulated host response to infection and identifies organ dysfunction as an acute increase in the Sequential Organ Failure Assessment (SOFA) score [6] of at least 2 points [7].

Sepsis research makes use of openly accessible ICU databases such as the single-centre database MIMIC-III [8] and eICU-CRD, which pools 208 US hospitals [9]. Studies on these databases range from observational analyses to prediction models [10], treatment-effect estimates [11] and comparisons of applied methods [12]. Generally, sepsis-related research depends not only on correctly identifying sepsis cases but also on precise dates of sepsis onset. The Sepsis-3 definition makes both possible, since the sepsis label and its onset can be computed directly from recorded clinical measurements. In contrast, diagnosis codes are unreliable, as sepsis codes in US hospitals identified only 32.3 % of patients with severe sepsis or septic shock (as defined prior to Sepsis-3) confirmed by medical record review [13].

However, the Sepsis-3 definition leaves many implementation details open, such as which measurements to use, over which time windows to evaluate them and how to handle missing values. Consequently, different implementations applied to the same data can yield substantially different sepsis labels. This variability is illustrated by our recent review, which covered 64 Sepsis-3 implementations on MIMIC-III and eICU-CRD [14]. Sepsis detection rates ranged from 3.4 % to 65.2 % in MIMIC-III and from 9.8 % to 47.9 % in eICU-CRD, even among studies with identical cohort definitions. Study characteristics such as the database, the unit of analysis and the infection detection method explained only about 9 % of this heterogeneity. Based on the publications and the source code analysed in the review, we grouped data preprocessing and label computation methods into six domains: parameter selection for each SOFA component (D1), temporal windows (D2), aggregation (D3), handling of missing values (D4), SOFA calculation (D5) and identification of suspected infection (D6). Most studies did not fully report how they implemented these six domains, and few published their source code. As a result, neither the effect of individual decisions nor the reproducibility of published rates could be assessed [14].

Recent studies have also shown that varying components of the Sepsis-3 definition itself, such as the infection criterion or the timing of onset, changes both labels and prediction performance [15, 16]. One of the studies also varied the imputation method and the SOFA version [16]. In contrast, parameter selection, temporal windows and aggregation have not yet been varied on identical data. To our knowledge, no study has compared single preprocessing decisions with a fixed reference, and no openly available pipeline exposes the decisions of domains D1–D5 as settings. A further gap concerns time, because labels are computed retrospectively from the complete record, whereas a real-time system only sees values that have already been documented.

To address these gaps, we developed medalabel, a pipeline that maps ICU data to the OMOP Common Data Model [17] and exposes the preprocessing decisions of domains D1–D5 as configurable settings. We hypothesized that single preprocessing decisions shift Sepsis-3 detection rates by an amount comparable to the variation between published studies, and that variants with similar rates label partly different patients. In addition, we examined whether reconstructions of published pipelines reproduce their published rates, whether the effects of decisions from different domains add up and how much the labels rely on values that a real-time system would not yet have.

## Materials and Methods

### Data Processing Pipeline

We developed medalabel (Figure 1), a configurable data processing pipeline based on the medallion architecture of data engineering, in which data pass through three layers of increasing quality called Bronze, Silver and Gold [18, 19]. It combines the preprocessing settings of published Sepsis-3 implementations in MIMIC-III and eICU-CRD in a single pipeline, so that each of them can be applied to either database. The pipeline thus serves as a test bench for label computation, in which individual decisions can be varied while data, cohort and definition remain fixed and the labelling pipelines of published studies can be reconstructed. Only the first layer is specific to a data source, and only the SOFA and label computation are specific to sepsis. Each layer ends with an automated profiling step for quality control. The technical specification is given in Supplement S1.

**Figure 1.**
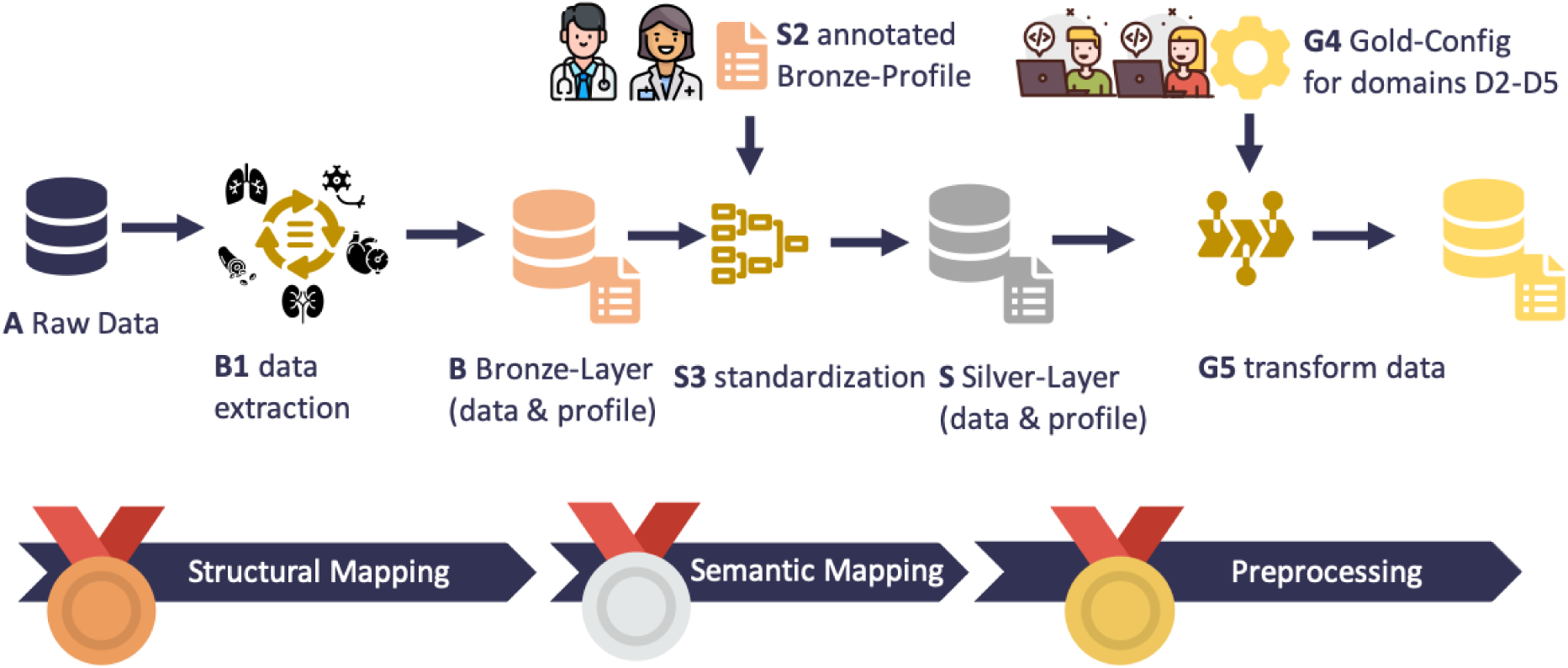
Data processing pipeline. In the structural mapping, raw ICU data (A) are extracted into the OMOP CDM (B1) and stored with their profile in the Bronze layer (B). In the semantic mapping, clinicians annotate the Bronze profile with OMOP concepts, units and plausibility ranges (S2), and the standardization step (S3) applies these annotations to produce the Silver layer (S). In the preprocessing, a configuration of the settings in domains D2–D5 (G4) determines how the data are transformed (G5) into SOFA scores and Sepsis-3 labels. The parameter selection (D1) follows from the annotated mapping (S2), from which each configuration reads the concepts of its parameter set. Expert input enters only at S2 and G4 (above the main path), and all other steps run automatically.

The Bronze layer maps the source data into the OMOP CDM v5.4 [17] without changing their content.

To simplify the following mapping to standard concepts, it fills the OMOP source_to_concept_map table with each source identifier, its name and, where the source data dictionary provides one, a standardized terminology code (LOINC, ICD-9, NDC). Bronze profiling calculates statistics for each source parameter, such as the number of observations, value distributions and measurement frequency, and reports them separately for each recorded unit (e.g., FiO_2_ in percent and as a fraction). These statistics show clinical experts what a source parameter contains, even where its name is ambiguous or its unit is missing. On this basis, the experts complete the mapping with standard OMOP concepts, unit rules, plausibility ranges and analytical tags, so that every standardization decision is clinically validated. The Silver layer applies this annotated mapping through the assignment of standard concepts, normalization of units and removal of implausible values.

The Gold layer selects the cohort, stores it in the OMOP cohort tables and retrieves only the concepts tagged for the analysis, which determines the parameter selection (D1). After normalizing vasopressor doses to body weight, deriving ventilation episodes and identifying suspected infection, the Gold layer applies four configurable stages, namely temporal aggregation by window size, step and per-parameter aggregation function (D2/D3), imputation of missing values (D4), SOFA subscore computation (D5) and Sepsis-3 label computation. Imputation proceeds in three levels of increasing uncertainty: derivation from related parameters by established clinical relationships (e.g., total GCS from its components), estimation from earlier values of the same stay (e.g., forward filling) and, where no information exists, constant default values (e.g., normal values). All settings are controlled through configuration files, so variants require no code changes.

On the technical side, the pipeline is started from a command line or scheduled with the workflow management software Apache Airflow, and it is packaged in Docker containers so that it runs unchanged on other systems. To keep test cycles short, the pipeline was developed on the MIMIC-III and eICU-CRD demo datasets and then applied unchanged to the full databases. More than 3,000 automated unit and integration tests (pytest test cases) check data integrity, transformation correctness and filter logic across all three layers.

### Data and Cohorts

In both databases, inclusion was restricted to adults (≥ 18 years) with an ICU stay of more than 24 h, and only the first ICU stay per patient was kept. For MIMIC-III (v1.4) [8], the analysis was further restricted to stays documented in MetaVision (iMDsoft), because microbiology cultures are underrepresented in the older CareVue system (Philips) [20]. The eICU-CRD (v2.0) [9] was used without further restriction. The primary outcome is the sepsis detection rate, i.e. the proportion of ICU stays labelled as septic. All times are counted from ICU admission (*t*_0_).

### Sepsis-3 Label and Reference Configuration

In the reference configuration, the pipeline aggregates the data of each ICU stay into 24 h windows that advance in 1 h steps (D2), takes the worst value of each parameter within a window, except that SpO_2_ is averaged and urine output is summed (D3), fills missing values with the three imputation levels described above (D4) and computes a SOFA score for every window (D5), giving one score per hour. A stay is labelled septic if its total SOFA score rises by at least 2 points above a baseline between 48 h before and 24 h after the onset of suspected infection [21]. The baseline is not a pre-illness value but the lowest total SOFA within the preceding 24 h. In MIMIC-III, suspected infection (D6) is defined as an antibiotic prescription combined with a microbiology culture obtained within 72 h before or 24 h after it, with onset at the earlier of the two [21]. We preferred this rule to ICD codes, which lack precise onset times and depend on coding practice [14]. In eICU-CRD, microbiology cultures are too sparse for this rule. Suspected infection is therefore taken from sepsis diagnosis records (ICD codes and the APACHE admission diagnosis), as in the two eICU-CRD repositories analysed, with onset at the time the diagnosis was entered. Because the two databases differ in how infection is identified, the comparison between them concerns the direction rather than the size of effects.

For parameter selection (D1), the reference combines the clinical parameters for SOFA calculation of the six public repositories analysed previously [14], four for MIMIC-III [11, 12, 22, 23] and two for eICUCRD [24, 25], into one union set (repositories in Table 2; parameter sets in Supplement S2, Section S2.1).

For MIMIC-III, we then searched the data dictionary for additional source codes of these parameters, and clinicians validated the resulting sets of both databases (Supplement S1, Section S1.3). Parameter selection (D1) was examined with the six repository-specific parameter sets, each restricted to the validated union (Supplement S2, Section S2.3). Infection identification (D6) was held fixed within each database.

Variants were derived from our previous analysis of the implementation decisions in the six repositories and the methods of the 64 reviewed studies [14]. Their consolidation and the decisions not modelled are described in Supplement S2 (Section S2.1). Each variant is labelled D*d*-S*n*, for domain *d* and setting *n*. In each domain, the reference is the method reported most often. The exception is the SOFA baseline, which few studies reported [14]. Its 24 h minimum follows [26], so that an acute deterioration can be detected at any hour of the stay. The Sepsis-3 consensus assumes a baseline of zero only for patients without known organ dysfunction [7]. With a zero baseline for all stays, patients with chronic organ dysfunction would meet the SOFA criterion throughout their stay, so this setting is examined as a variant (D5-S15) and not used as the reference.

### Study Design

The analyses were organized in three tiers and an analysis of information time, each answering one question (Table 1). Every configuration is a complete pipeline run in which the Gold layer recomputes aggregation, imputation, SOFA scores and labels for every hour of every ICU stay, which yields about 0.5 million hourly rows per configuration in MIMIC-III and 6.9 million in eICU-CRD (computational resources in Supplement S1, Section S1.1).

**Table 1.** Study design. All configurations run on the same Silver data. Except for Tier 2, which uses the cohorts of the repositories, all analyses share one fixed cohort per database.

| Analysis | Question | What is varied |
|---|---|---|
| Tier 1 | How much does each decision shift the label on its own? | 51 variants that each change one setting (D1–D5), all others at the reference |
| Tier 2 | Can published labels be reproduced? | 6 reconstructions of complete repository pipelines, including their own cohorts (4 in MIMIC-III, 2 in eICU-CRD) |
| Tier 3 | Do effects of different domains combine additively? | The single-setting variant with the largest Tier 1 effect from each of the domains D2–D5, combined in all 6 possible pairs, in each database |
| Information time | How much do labels rely on values not yet documented? | Values available from their documentation time instead of their measurement time (MIMIC-III) |

#### Tier 1

The 51 variants comprise 6 parameter sets (D1), 10 temporal windows (D2), 3 aggregation rules (D3), 11 settings for missing values (D4) and 21 settings of the SOFA calculation (D5).

#### Tier 2

The pipelines of the six repositories listed above were reconstructed from their source code by mapping their decisions to the corresponding pipeline settings, including their parameter sets and cohort definitions. Because some decisions cannot be represented in the pipeline (Supplement S2, Section S2.1), the reconstructions are approximations. Their rates were compared with the published ones.

#### Tier 3

Effects combine additively if two variants applied together shift the rate by the sum of their single effects. For each of the domains D2–D5, we selected the variant with the largest Tier 1 effect in MIMIC-III, excluding bounds and variants that change several settings at once, so that each factor represents one decision. For comparability, the same four variants were combined in all six pairs in both databases (Figure 3).

#### Information time

Like all repositories analysed, every configuration uses each value from the time it was measured (charttime). For bedside observations, MIMIC-III also records when a value was entered into the clinical information system and thus became available (storetime). Because the OMOP CDM holds a single timestamp per event [27], we extended its measurement and drug exposure tables with a storetime column and re-ran the reference with documentation-time availability, in which each value counts at its charttime but only once it has been documented (Supplement S1, Section S1.2, and Supplement S2, Section S2.4). The contrast is restricted to MIMIC-III because eICU-CRD records no storetime. Within MIMIC-III, results from the laboratory and microbiology tables carry none either, except laboratory values that are also charted at the bedside. The contrast therefore underestimates how many values a real-time system would not yet have.

#### Further contrasts

Supplement S2 (Section S2.4) reports three additional settings, namely the first measurement instead of ICU admission as time origin, the inclusion of CareVue stays in MIMIC-III and the stricter infection rule.

### Statistical Analysis

The effect of each variant was expressed as the signed difference between its detection rate and that of the reference in percentage points, and each domain was characterized by the difference between the highest and the lowest rate among its variants. Two variants that do not correspond to a published method, the best-case aggregation (D3-S4) and the additional variant D2-S10, were reported but not counted in this difference (rationale in Supplement S2, Section S2.1). Because all configurations run on the same fixed cohort, differences between them are exact and were compared without significance tests. Because the cohorts are large, the sampling uncertainty of each rate is small (95 % Wilson confidence interval within ±0.9 pp of the rate), so confidence intervals are not shown for individual rates. Shifts in SOFA total and subscores were summarized as mean differences from the reference (Supplement S2, Section S2.3).

Whether two configurations labelled the same stays was quantified with the Jaccard index, which is the number of stays labelled septic by both divided by the number labelled septic by at least one. It is 1 when both label exactly the same stays and 0 when they share none, independent of whether the rates differ. The index alone does not show whether a variant adds septic stays, removes them or exchanges them. For every variant, Supplement S2 (Section S2.3) therefore also lists how many stays gained and how many lost the label compared with the reference. Tier 3 interactions were computed as the observed effect of a combination minus the sum of the two single effects, in percentage points. An interaction of 0 pp means that the effects simply add. As an alternative, we compared the observed rate with the product of the relative changes of the two settings. Agreement between databases was described with the Spearman rank correlation of the D2–D5 variant rates and with the direction of variants that shifted the rate by at least 1 pp in both. Tier 2 rates were compared with the published rates as absolute differences, and all rates with the range of published rates [14]. Analyses used Python with pandas and SciPy.

## Results

After applying the inclusion criteria, the reference cohorts comprised 13,388 ICU stays in MIMIC-III and 94,782 in eICU-CRD, with reference detection rates of 31.5 % and 8.2 %, respectively. The catalogue of the 51 Tier 1 variants is provided in Supplement S2 (Section S2.1).

### Source Data

Before preprocessing, the Bronze layer held 21,813 source codes with about 365 million records in MIMIC-III, of which 349 codes were mapped to 39 OMOP concepts for the Sepsis-3 label. In eICU-CRD, 255 of 14,276 codes with about 985 million records were mapped to 27 concepts. Both databases recorded the same parameter under several names, codes and unit spellings. In MIMIC-III, urine output had to be determined from 12 source codes named after the collection route (e.g., “Foley” and “Ileoconduit”). Mechanical ventilation was derived from 78 source codes. In eICU-CRD, norepinephrine appeared under 19 names, some of which include the concentration, and a single platelet code carried 55 spellings of its unit. Within an ICU stay, MIMIC-III recorded vital signs about hourly and laboratory values every 8 to 24 h on average, whereas eICU-CRD recorded SpO_2_ about every 6 min and mean arterial pressure every 15 min, but laboratory values only every 15 to 37 h (Supplement S2, Section S2.2).

### Single Decisions (Tier 1)

Detection rates varied widely across preprocessing configurations (Figure 2). In MIMIC-III, Tier 1 rates ranged from 15.3 % without imputation to 58.6 % with a zero SOFA baseline (reference 31.5 %). Rates varied most within SOFA calculation, where the highest and the lowest rate differed by 38.6 pp (zero baseline D5-S15 against 1 h baseline window D5-S18), followed by missing values (20.3 pp, cohort-mean fill D4-S10 against no imputation D4-S12), temporal windows (15.4 pp, 1 h windows D2-S2 against first 24 h only D2-S8) and aggregation (10.1 pp, reference against mean aggregation D3-S2).

**Figure 2.**
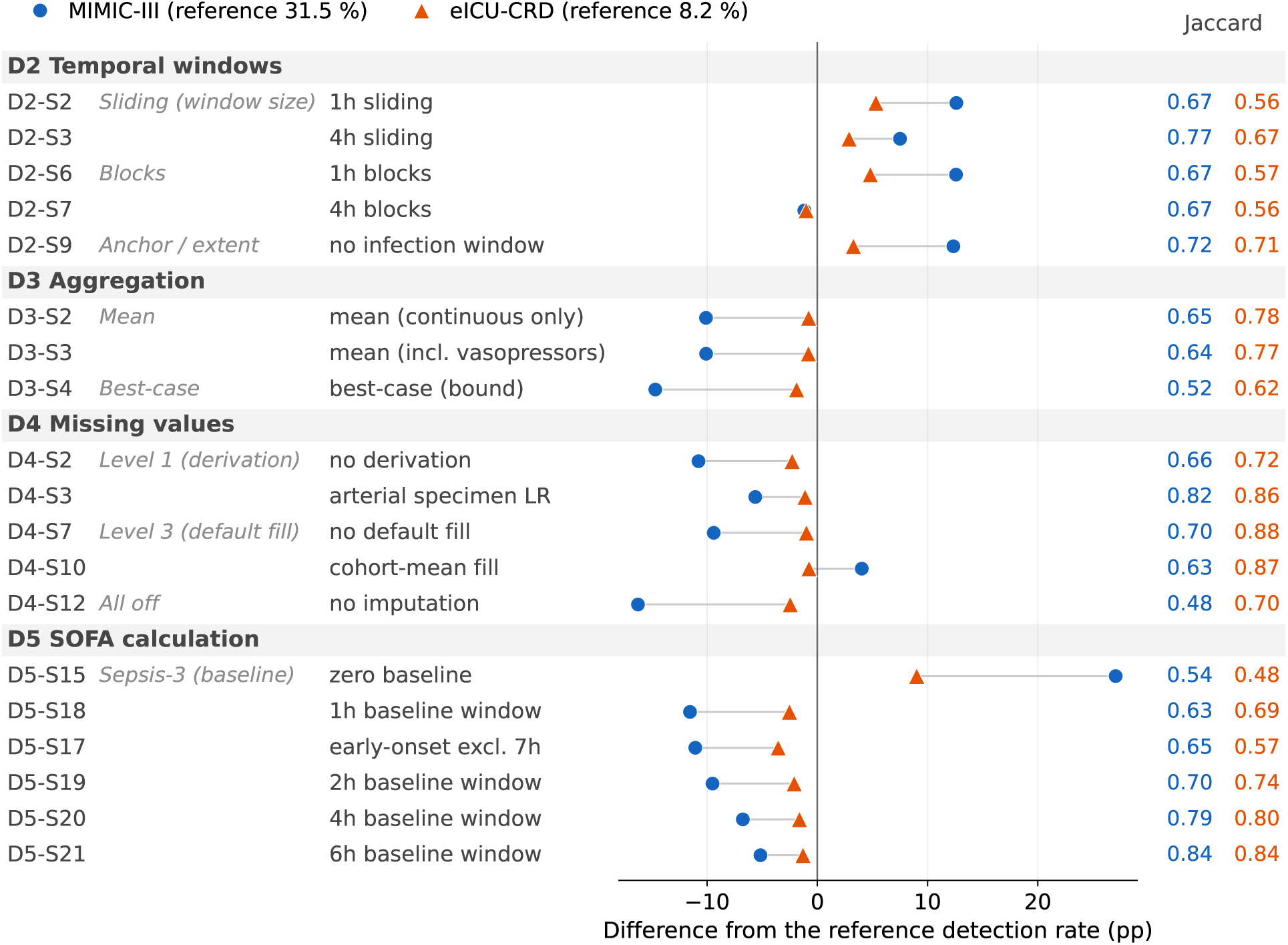
Effects of selected Tier 1 variants on the detection rate, as the difference from the reference in percentage points (MIMIC-III 31.5 %, eICU-CRD 8.2 %). Shown are the 19 variants of the processing domains D2–D5 that, in at least one database, shifted the rate by at least 5 pp or labelled septic stays with a Jaccard index below 0.70 against the reference. Rows are grouped by domain and subgroup and sorted by the MIMIC-III effect within each subgroup; D3-S4 is a lower bound and not a published method. The right-hand columns give the Jaccard index in MIMIC-III (blue) and eICU-CRD (orange).

Within SOFA calculation, the baseline of the Sepsis-3 criterion dominated. Counting every total SOFA of 2 or more as an increase (zero baseline, D5-S15; Figure 2) raised the rate by 27.1 pp, and shortening the 24 h window in which the baseline is sought lowered it step by step, from −2.3 pp at 12 h to −11.6 pp at 1 h. Excluding cases with sepsis onset within the first 7 h after ICU admission (early-onset exclusion, D5-S17) lowered it by 11.1 pp, whereas no variant of organ scoring moved it by more than 2.9 pp (all D5 variants and their effect on each SOFA subscore in Supplement S2, Section S2.3). In the other domains, the largest reductions came from disabling imputation altogether (D4-S12, −16.3 pp) or only the derivation of missing values from related parameters (D4-S2, −10.8 pp) and from mean instead of worst-value aggregation (D3-S2, −10.1 pp). For every imputation variant, Supplement S2 (Section S2.3) shows how many values each of the three imputation levels fills. The largest increases came from 1 h windows (sliding windows D2-S2 and non-overlapping blocks D2-S6, +12.6 pp each) and from scoring the SOFA increase over the whole stay instead of the window around suspected infection (D2-S9, +12.3 pp).

In eICU-CRD, rates were less sensitive to preprocessing. Tier 1 rates ranged from 4.6 % with the earlyonset exclusion to 17.2 % with a zero baseline (reference 8.2 %). As in MIMIC-III, the highest and the lowest rate differed most within SOFA calculation (12.6 pp, zero baseline D5-S15 against early-onset exclusion D5-S17), followed by temporal windows (6.3 pp, 1 h windows D2-S2 against 4 h blocks D2-S7), missing values (5.6 pp, unmeasured urine output scored as anuria D4-S11 against no imputation D4-S12) and aggregation (0.8 pp, reference against mean aggregation D3-S2). One variant, the scoring of renal replacement therapy (D5-S10), was run only in MIMIC-III. Across the remaining 44 D2–D5 variants, a variant that yielded a higher detection rate in MIMIC-III usually also yielded a higher rate in eICU-CRD (Spearman rank correlation of the 44 pairs of rates, *ρ* = 0.90), and every variant that shifted the rate by at least 1 pp in both databases did so in the same direction (16 of 16).

Most variants relabelled only a small part of the stays. The median Jaccard index between a variant’s septic stays and those of the reference was 0.90 in MIMIC-III and 0.88 in eICU-CRD. The septic stays changed most when imputation was disabled (0.48 in MIMIC-III), with best-value aggregation (0.52 in MIMIC-III; D3-S4, a lower bound and not a published method) and with a zero baseline (0.54 in MIMIC-III, 0.48 in eICU-CRD). Variants with similar rates could still label different patients. Non-overlapping 4 h blocks, the time steps of one repository (Table 2, No. 4), changed the MIMIC-III rate by only −1.2 pp, yet 748 stays gained and 906 lost the label (Jaccard 0.67). In eICU-CRD the rate changed by −1.0 pp while 1,560 stays gained and 2,514 lost it (Jaccard 0.56).

**Table 2.** Tier 2 reconstructions of the six repository pipelines, each with its own cohort definition. Rates in %, deviation in percentage points, computed from the unrounded rates. *^a^*With the repository’s own antibiotic list, which selects drugs by GSN code; as the source data in the pipeline carry no GSN code, it was approximated by the drug name and route pairs whose prescriptions carry one of these codes in at least half of the cases. *^b^*Rate or cohort size inferred from the publication rather than reported [14]. *^c^*Reconstructed with the diagnosis-based identification of the repository [14]; in brackets with the SOFA criterion additionally required.

| **No.** | **Repository** | **Database** | **ICU stays** | **Reconstructed** | **Published** | **Deviation** |
|---|---|---|---|---|---|---|
| 1 | sepsis3-mimic [12] | MIMIC-III | 14,430 | 43.4 | 49.1 | −5.7 |
| 2 | MIMIC-Code [22] | MIMIC-III | 59,936 | 29.7 | – | – |
| 3 | sepsis-3-labels [23] | MIMIC-III | 52,899 | 17.0 | 14.9<sup>b</sup> | +2.2 |
| 4 | AI Clinician [11] | MIMIC-III | 50,003 | 30.6<sup>a</sup> | 27.8<sup>b</sup> | +2.8 |
| 5 | AI-enrichment [24] | eICU-CRD | 18,064 | 25.9<sup>c</sup> (23.4) | 16.4<sup>b</sup> | +9.5 |
| 6 | trajPLT [25] | eICU-CRD | 13,764 | 20.8<sup>c</sup> (16.8) | – | – |

For parameter selection (D1), only the parameter set of the reference was replaced by that of a repository, while all other settings and the cohort stayed at the reference. These parameter sets, restricted to the clinician-validated union, had little effect (range 4.8 pp in MIMIC-III and 1.1 pp in eICU-CRD). Three MIMIC-III sets (Table 2, No. 1, 2 and 3) and one eICU-CRD set (No. 6) labelled virtually the same stays as the reference (Jaccard 1.00). The parameter set of repository No. 4 lowered the MIMIC-III rate by 4.8 pp and that of No. 5 lowered the eICU-CRD rate by 1.1 pp (Jaccard 0.85 and 0.87). Both labelled a subset of the reference stays, with 649 and 1,029 stays losing the label and 3 gaining it in each database (Supplement S2, Section S2.3).

At the score level, the largest shifts in mean total SOFA in MIMIC-III came from filling missing values with the cohort mean (D4-S10, +2.9 points), scoring the whole stay (D2-S9, +2.3) and disabling imputation (D4-S12, −2.0). The CNS and urine output variants changed only the CNS and the renal subscore, respectively, and the variants of Sepsis-3 onset and baseline changed detection rates without altering any subscore (Supplement S2, Section S2.3).

### Published Pipelines (Tier 2)

The four reconstructions with a published rate deviated from it by between −5.7 and +9.5 pp (Table 2), although each was run on the same database version as the original study. In eICU-CRD, requiring the SOFA criterion in addition to the repositories’ diagnosis codes lowered the reconstructed rates by 2.5 and 4.0 pp. Because each reconstruction keeps the cohort definition of its repository, Tier 2 rates are not directly comparable with the fixed-cohort Tier 1 rates. Across Tier 1 and Tier 2, the rates ranged from 15.3 % to 58.6 % in MIMIC-III and from 4.6 % to 25.9 % in eICU-CRD.

### Combined Decisions (Tier 3)

Tier 3 crossed 1 h blocks (D2-S6), mean aggregation (D3-S2), no derivation (D4-S2) and a zero baseline (D5-S15) in all six pairs (Figure 3; Supplement S2, Section S2.3). In MIMIC-III, only the combination of no derivation and zero baseline (D4-S2 with D5-S15) came close to the sum of the two single effects (interaction +0.9 pp), whereas the other five pairs deviated by 4.4 to 11.8 pp. In these pairs, one setting dominated and the other lost much of its effect. The 1 h blocks combined with mean aggregation (D2-S6 with D3-S2) labelled almost the same stays as the 1 h blocks alone (Jaccard 0.99), and the zero baseline combined with mean aggregation (D5-S15 with D3-S2) labelled almost the same stays as the zero baseline alone (Jaccard 0.94). Conversely, the 1 h blocks, which raised the rate by 12.6 pp on their own, added less than 1 pp when no derivation or the zero baseline was applied as well (D2-S6 with D4-S2 and with D5-S15). The stays they added were almost all labelled by the zero baseline already (92 % in MIMIC-III, 99 % in eICU-CRD), and without derivation only 8 % and 15 % of them remained septic. A multiplicative model, in which the relative changes of the two settings are multiplied, did not fit better. It came closer to the observed rate than the additive model in two of the six pairs in each database and deviated by up to 22.4 pp in MIMIC-III and 10.1 pp in eICU-CRD.

**Figure 3.**
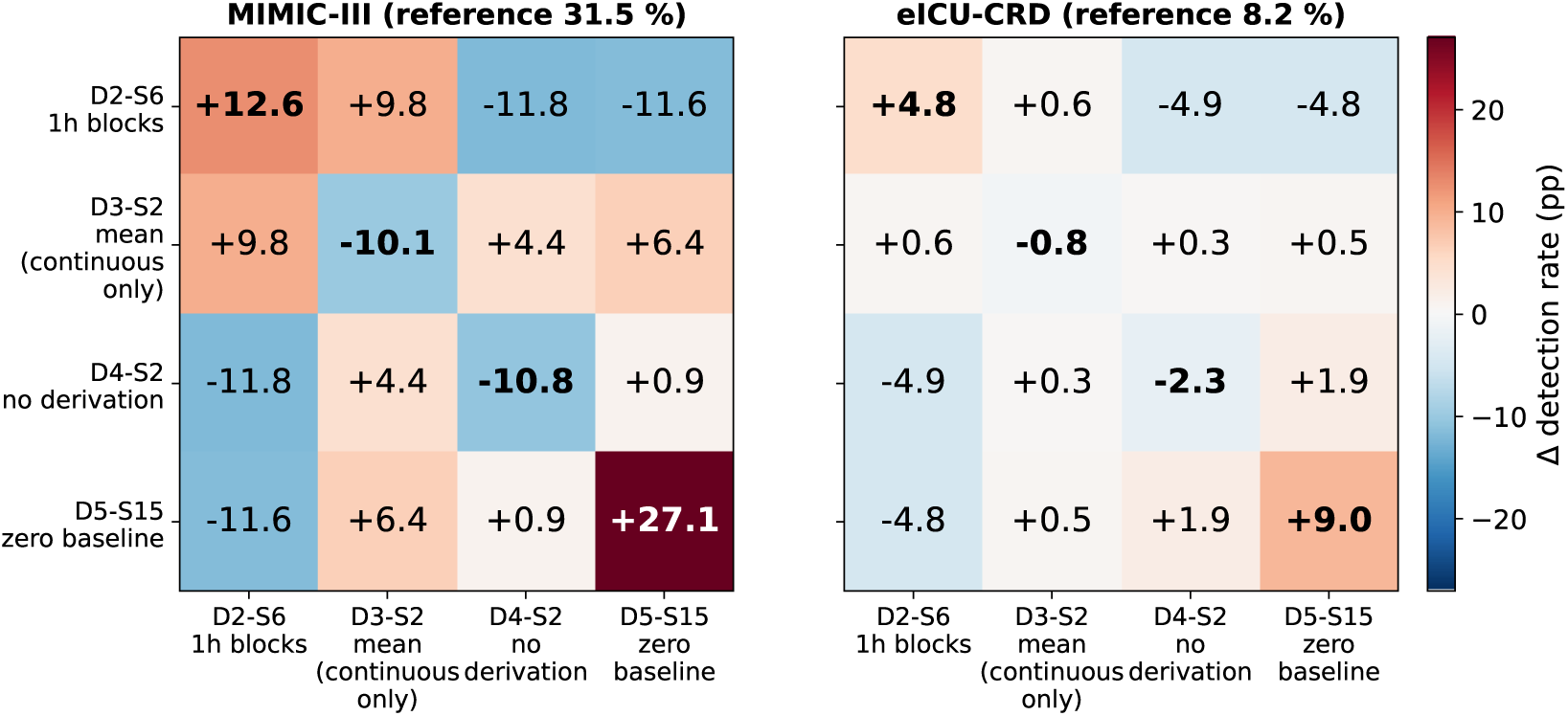
Tier 3 interaction analysis. Diagonal cells (bold) show the single effect of each variant alone, as the deviation from the reference rate in percentage points; off-diagonal cells show the two-way interaction, i.e. the observed effect of the combination minus the sum of both single effects. Red cells raise and blue cells lower the rate; white means no deviation, i.e. for interactions that the effects simply add. Because all configurations run on the same cohort, the values are exact differences rather than estimates, and no significance test applies. Both panels share one colour scale.

In eICU-CRD, the pattern was the same on a smaller scale. All four single effects and all six interactions had the same sign as in MIMIC-III, and the zero baseline again had the largest single effect (+9.0 pp). Only the two pairs of the 1 h blocks with no derivation and with the zero baseline deviated by more than 2 pp (−4.9 and −4.8 pp).

### Information Time and Further Contrasts

Using each value only once it was documented, as a real-time system would, lowered the MIMIC-III rate from 31.5 % to 26.5 % (−5.0 pp). Of the 4,223 septic stays of the reference, 918 (21.7 %) lost the label and 246 other stays gained it, so that both labels shared 3,305 stays (Jaccard 0.74). In these shared stays, the label was first met at the same hour in 1,726 (52 %), later in 1,481 (45 %) and earlier in 98 (3 %). Where it was met later, the median delay was 3 h, and in 415 stays it was at least 6 h. On average, documentation followed measurement by 14–40 min for most bedside parameters and by 55–129 min for laboratory values that are also charted at the bedside (Supplement S2, Section S2.4).

Of the further contrasts (Supplement S2, Section S2.4), anchoring *t*_0_ at the first measurement instead of ICU admission changed rates by at most 0.9 pp in MIMIC-III and 2.2 pp in eICU-CRD, including CareVue stays lowered the MIMIC-III rate to 26.2 % (CareVue alone: 22.4 %), and the stricter infection rule lowered the reference rate of 31.5 % to 19.1 %.

## Discussion

With data, cohort and Sepsis-3 definition held fixed, single preprocessing decisions shifted the detection rate from 31.5 % to between 15.3 % and 58.6 % in MIMIC-III and from 8.2 % to between 4.6 % and 17.2 % in eICU-CRD. Variants with similar rates labelled partly different patients, reconstructions of published pipelines deviated from the published rates, the effects of two decisions rarely added up, and labels computed from measurement times relied on values that were documented only later. A considerable part of the variation between published studies can therefore arise from the preprocessing decisions of domains D1–D5 alone, even with the infection criterion held fixed, which supports our first hypothesis.

Together with the reconstructions of published pipelines, which reached 25.9 % in eICU-CRD, our rates covered 70 % of the range published for MIMIC-III (3.4–65.2 %) and 42 % of that for eICU-CRD (9.8– 47.9 %) [14]. These percentages compare only the lowest and the highest rate on each side. In our analyses, these extremes come from single variants, in MIMIC-III from disabling imputation and from the zero baseline, and in eICU-CRD the highest rate comes from a reconstruction with its own cohort. They should therefore not be read as the share of the published differences that preprocessing explains. They show that preprocessing alone can move the rate across a similarly wide range. The remaining range stems from studies that differ in what our design holds fixed, such as additional cohort filters, admissions instead of ICU stays as the unit of analysis, or an unreported infection definition [14], and from implementation details that our 51 variants do not cover.

The decisions that mattered most are rarely reported [14]. In MIMIC-III, the baseline of the SOFA increase, the handling of missing values, the aggregation within a window, the window granularity, the restriction to the window around suspected infection and the exclusion of early-onset cases each shifted the rate by more than 10 pp, whereas the clinician-validated repository parameter sets mattered little (at most 4.8 pp). In percentage points, eICU-CRD was less sensitive, but its reference rate is also lower.

Relative to the reference rate, the zero baseline and the 1 h windows had larger effects in eICU-CRD than in MIMIC-III (+110 % against +86 % and +65 % against +40 %), whereas disabling imputation and mean aggregation had smaller ones (−30 % against −52 % and −10 % against −32 %). The direction of effects agreed between the databases whenever a variant mattered in both, but effect sizes did not transfer one-to-one. Effects of decisions from different domains rarely combined additively. When one setting dominated, such as the zero baseline or 1 h blocks, the other lost much of its effect. This is partly inherent to the scale, because a stay can be labelled septic only once and two settings that act on the same stays cannot add their effects. Effects of individual decisions therefore cannot simply be summed.

Our reconstructions approximate the published pipelines with the settings available in our pipeline. Run on the same database version as the original studies, they yielded rates that deviated from the published ones by 2.2–9.5 pp in both directions, and these deviations cannot be attributed to a single cause. Part of these deviations lies in the comparison itself. Only one repository (Table 2, No. 1) reports both the number of septic cases and the cohort size, whereas for the other three the denominator had to be inferred from the size of the database [14], which tends to understate the rate when the published cohort was filtered further. Yet even this repository, with both numbers reported, deviated by 5.7 pp. The reconstructions also cover most, but not all, implementation decisions of the repositories (Supplement S2, Section S2.1), and published rates further depend on undocumented choices, software versions and extraction details that the code does not capture.

These results add a layer to known reproducibility problems. Machine learning for health suffers from persistent reproducibility gaps [28, 29], and systematic reviews of sepsis prediction could not pool performance across studies because of varying sepsis definitions [30, 31]. Open ICU databases record no reliable sepsis timestamps [32], and implementing Sepsis-3 in AmsterdamUMCdb raised the same operational questions [33], so every label has to be reconstructed by preprocessing. Our results indicate that this reconstruction is itself a source of variability. Varying the definition itself changes labels and predictive performance [15]. Closest to our work, Mackensen et al. [16] crossed 2,520 variants of the Sepsis-3 definition and attributed most label variability to the suspected-infection criterion, with imputation and window length contributing little. Where the two designs overlap, the results agree, as adding the four-day antibiotic rule of the CDC Adult Sepsis Event [13] lowered our MIMIC-III reference rate from 31.5 % to 19.1 %. They diverge on imputation, which contributed little in their design but shifted our rates by up to 16.3 pp when disabled, and on evaluating the SOFA increase over the whole stay, which their design did not include and which shifted our rates by 12.3 pp. Harmonization frameworks such as ricu [34] provide a harmonized implementation across databases, whereas our pipeline exposes the decisions behind it, so that their effect can be measured.

In line with our second hypothesis, similar detection rates did not imply that the same patients were labelled. Most variants relabelled only a small part of the stays, but some replaced about a fifth of the septic stays while leaving the rate almost unchanged, and the subscore analyses showed that variants act on specific organ systems. Two studies with similar rates may therefore identify sepsis through different organ dysfunction patterns in partly different patients. For prediction models, this may limit the comparability of reported performance and the transfer of published models to practice. In observational and treatment-effect studies, the label decides which patients are analysed as septic, so estimates of incidence, outcome and treatment effect may inherit the same differences.

Every published label we examined, and our reference, also uses each value from its measurement time, as if it were available the instant it was recorded. Counting values only once documented lowered the MIMIC-III rate by 5.0 pp. About one in five septic stays of the reference lost the label, and in almost half of the remaining ones it was met later, so these labels depend on values that a real-time system would not yet have. Because results from the laboratory table carry no documentation time, this is a lower bound. A related problem was observed after deployment, where a widely implemented proprietary sepsis model reached an AUROC of 0.63 in external validation [35] and 0.47 once predictions made after clinical recognition were excluded [36]. Prospective evaluation can succeed [37, 38], but its results transfer only if the label pipeline is reported as precisely as the model.

The Lancet Commission on Sepsis counts the lack of consensus on definitions and surveillance criteria among the central problems of the field [39]. Our results suggest that even a consensus definition does not yield consistent labels unless its operationalization is agreed and reported. A common data model is necessary but not sufficient, because identical OMOP schemas can hold semantically different content [27] and identical data still yield different labels. Mapping data into the model is itself demanding. Even within one hospital, the same clinical concept was recorded under many source codes, and the 208 hospitals of eICU-CRD added local names, unit spellings and dosing conventions. Such heterogeneity can be expected to grow with every additional site, and recording the mapping in a reviewable file, as in the Silver layer, makes this step transparent and reusable.

Labels would become comparable if studies reported at least the SOFA baseline, the handling of missing values, the aggregation rule, the window and observation period, the handling of early-onset cases, the information time and the parameter set of each SOFA component [14]. Reporting guidelines such as TRIPOD [40], STROBE [41] and CONSORT [42] could cover these decisions for database-derived labels. For deployed models, these decisions also belong in the documentation that reaches clinicians, for which clinician-facing model cards offer a place [43]. How such information is presented matters as well, because the way predictive uncertainty is shown in sepsis decision support changed physicians’ advice uptake, accuracy and trust [44]. Publishing the preprocessing code with the article and testing the decisions with the largest effects identified here in sensitivity analyses, ideally in more than one database, would allow readers to judge how robust a result is. Configurable pipelines make such analyses feasible without rewriting code. The underlying architecture is not specific to sepsis and has a low entry barrier. In a master’s course, 11 student groups each built a simplified medallion pipeline on the MIMIC-IV demo dataset for a disease-specific research question [45].

In a follow-up study, we plan to apply the pipeline to the ICU data of our institution, whose extraction infrastructure is in place [46]. Because the Gold layer reads only the standardized OMOP representation, the analyses can run on institutional data without their developers accessing it, following the code-to-data model of OMOP network studies [47] that avoids the barriers to sharing routine clinical data [48, 49]. There, we intend to compare our configurations with sepsis diagnoses and onset times adjudicated by intensivists, extending comparisons of Sepsis-3 definitions with expert labels [16, 50] to the preprocessing that precedes the definition. Where structured infection records are sparse, sepsis-related features extracted from clinical notes [51] could complement the infection criterion. The clinician-validated annotations of this study can further serve as a reference for automated concept mapping, where large language models still require expert review [52].

### Limitations

Both databases originate from the US, and validation in other healthcare systems remains to be done. The variants form a structured sample of the decision space rather than a complete enumeration. Parameter selection was limited to the six repository sets, infection identification was held fixed apart from one contrast, and Tier 3 combined four selected variants in pairs only, without higher-order combinations. All effects are relative to the reference, whose 24 h minimum baseline is not the most common choice. Tier 3 indicates how they change with the zero baseline as starting point, which most of the repositories use [14]. No derivation still lowered the rate by 9.9 pp and mean aggregation by 3.7 pp, whereas the 1 h blocks added only 0.9 pp. The direction of the effects was thus unchanged, but their size depends on the reference. Because cultures are too sparse in eICU-CRD, suspected infection had to be taken from diagnosis records there, although such records miss many septic patients [13]. The eICU-CRD rates are therefore likely to underestimate sepsis, absolute rates are not comparable between the databases, and eICU-CRD serves to test whether the direction of the effects is replicated. The eICU-CRD reference rate of 8.2 % also lies below the lowest published rate, for which we see two reasons. The reference requires a SOFA increase in addition to the diagnosis-based infection criterion, and in our reconstructions of the two eICU-CRD repositories this requirement lowered the rates by 2.5 and 4.0 pp. The cohort differs as well, because the reference is restricted to first ICU stays of more than 24 h, and under the same requirement the cohorts of the two repositories yielded 16.8 % and 23.4 %. The documentation-time contrast is restricted to MIMIC-III and excludes results from the laboratory table. When PaO_2_ is missing, the respiratory subscore depends on the chosen SpO_2_/FiO_2_ equivalence, of which we tested one alternative [53] but not others [54, 55]. Results refer to the original SOFA score, and SOFA-2 [56] will add further decisions, which the pipeline can represent through configuration. Finally, our analyses show how much preprocessing changes the label, not which label is clinically correct, because neither database provides a clinical reference standard.

## Conclusion

With data, cohort and infection criterion held fixed, the preprocessing decisions of domains D1–D5 changed how many and which patients were labelled septic by Sepsis-3 in two ICU databases. The SOFA baseline had the largest effect in both databases, and the handling of missing values and the temporal window followed. The effects of two decisions rarely added up, and in MIMIC-III retrospective labels relied on values that a real-time system would not yet have. Sepsis-3 labels are therefore comparable only if their preprocessing is known.

We advocate routine sensitivity analyses of preprocessing and standardized reporting of the implementation decisions in domains D1–D6. The configurable pipeline presented here makes these decisions explicit. Which of the resulting labels is clinically correct remains to be tested against clinician-adjudicated cases.

## Supporting information

Supplement S1: Data Processing Pipeline

Supplement S2: Configurations and Detailed Results

## Ethics Statement

MIMIC-III and eICU-CRD contain de-identified data. The use of MIMIC-III for research was approved by the institutional review boards of the Massachusetts Institute of Technology and Beth Israel Deaconess Medical Center with a waiver of informed consent, and analyses of eICU-CRD are exempt from institutional review board approval. Both databases were accessed under their PhysioNet data use agreements.

## Use of AI Tools

Claude Code (Anthropic; Opus 4.6–5.5, Fable 5.2) was used to assist with (1) developing and testing the code of the pipeline and of the analysis, (2) preparing figures and tables, and (3) language editing of the manuscript and the supplements. All AI-generated outputs were verified by the authors.

## Data Availability

MIMIC-III and eICU-CRD are available from PhysioNet to credentialed users. The pipeline code, its configurations and the analysis code will be made publicly available upon publication.

## Acknowledgements

This work was supported by the de.NBI Cloud within the German Network for Bioinformatics Infrastructure (de.NBI) and ELIXIR-DE (Forschungszentrum Jülich and W-de.NBI-001, W-de.NBI-004, W-de.NBI-008, W-de.NBI-010, W-de.NBI-013, W-de.NBI-014, W-de.NBI-016, W-de.NBI-022).

