## Supplement S1: Data Processing Pipeline for "Effect of Data Preprocessing Decisions on Sepsis-3 Labels in MIMIC-III and eICU-CRD: A Sensitivity Analysis"

This supplement provides technical details on the data processing pipeline described in the main text, including OMOP CDM adaptations, layer-specific processing steps, deployment architecture, and the clinical parameter identification workflow.

### S1.1 System Architecture

The `medalabel` pipeline follows the three-layer medallion architecture [1, 2]. Its core processing logic is a standalone Python application (`src/` modules) without dependency on workflow orchestration software and can be run from an interactive command-line interface. The configurations of this study were executed by a configuration runner that processes each configuration (pipeline, profiling and summarization) in a separate process, with configurations running in parallel. For routine operation, each layer is additionally available as a directed acyclic graph (DAG) in Apache Airflow 3. A DAG defines processing steps (tasks) and their dependencies, so that steps run in the correct order. Airflow adds per-task retries with exponential backoff (three retries; 2–30 min for Bronze and Silver, 5–60 min for Gold), progress monitoring through a web interface and parallel execution of independent tasks.

Three DAGs are defined. The Bronze DAG (18 tasks, currently for MIMIC-III only) first creates the schema. It then fills the OMOP tables for patients, hospital admissions and ICU stays (`person`, `visit_occurrence`, `visit_detail`) one after another, because each of them refers to the one before it. The remaining ten source tables are mapped in parallel, followed by the creation of constraints, validation and profiling. The Silver DAG (12 tasks) runs the Silver steps and the profiling in sequence. The Gold DAG prepares the result table, processes the ICU stays in parallel chunks (five by default, configurable with `performance.max_chunks`), creates the indices and runs the profiling.

The system is containerized as a Docker Compose stack with two services: a PostgreSQL instance for Airflow metadata and an Airflow 3 standalone container (webserver, scheduler and triggerer) that mounts the source code, configuration and data directories. The clinical data reside in a separate PostgreSQL instance. Figure 1 shows the deployment.

*Computational resources.* All configurations ran on a de.NBI Cloud virtual machine with 28 virtual cores, 256 GB RAM and 3.5 TB storage, which hosts the MIMIC-III and eICU-CRD PostgreSQL databases in Docker containers. Each configuration is a complete Gold run and produces about 0.5 million hourly rows in MIMIC-III and 6.9 million in eICU-CRD. Across Tiers 1–3 and the further settings, 135 configurations were run on the full databases, up to 20 of them in parallel.

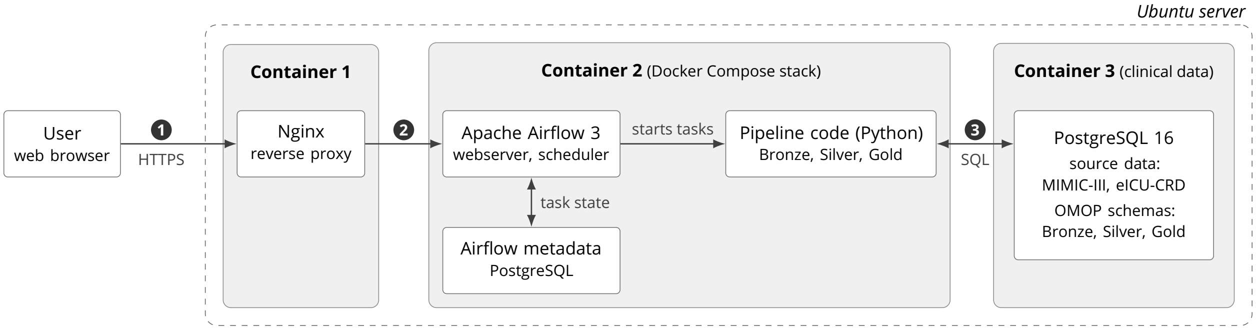


**Figure 1.** Deployment architecture on an Ubuntu server, numbered from left to right. Container 1 is an Nginx reverse proxy, operated outside the stack, that provides remote access to the Airflow web interface. Container 2 runs the repository's Docker Compose stack with Apache Airflow 3, the Python pipeline code and the Airflow metadata database. Container 3 is a separate PostgreSQL 16 instance that holds the source data and the OMOP schemas of the three layers. Numbered flows: (1) users reach the Airflow interface via HTTPS, (2) Nginx forwards these requests to the Airflow webserver, (3) the pipeline code reads from and writes to the clinical database.

**Table 1.** Deployment stack components.

| Component | Technology | Purpose |
|---|---|---|
| Orchestration | Apache Airflow 3 | Workflow management, scheduling, monitoring |
| Clinical data | PostgreSQL 16 | Source and OMOP data (MIMIC-III, eICU-CRD); separate instance |
| Metadata DB | PostgreSQL | Airflow metadata and task state |
| Containerization | Docker Compose | Reproducible deployment |
| Language | Python 3.11+ | Core processing logic |

### S1.2 OMOP CDM v5.4 Adaptations

We instantiated the official OMOP CDM v5.4 schema with foreign key constraints between populated tables. Several adaptations accommodate ICU time-series data and multi-source ingestion.

*Storetime columns.* The OMOP CDM does not distinguish between the time a clinical event occurred and the time it was documented. For real-time use this distinction matters, because a value is available only once documented. In the MIMIC-III demo data, 12.7 % of charted values carry a `storetime` before their charttime. Such values are not affected by the documentation-time setting, because they are already documented at their charttime. We added the columns measurement_storetime and drug_exposure_storetime. In MIMIC-III, results from the laboratory and microbiology tables have no storetime, so their measurement time is used instead. Laboratory values that are also charted at the bedside carry the storetime of that entry. In eICU-CRD, storetime is set to the measurement time. The Gold layer uses measurement time by default and can additionally require documentation before a value enters a window (Supplement S2, Section S2.4).

*Mapping table extensions.* The source_to_concept_map table was extended with the columns target_concept_code, source_table, source_unit, unit_corrected, target_unit, unit_concept_id, conversion, valid_low, valid_high and tags (PostgreSQL TEXT[] array). Parameters excluded during annotation are not imported. Tags such as sepsis3_mimic, weight and infection let the Gold layer load only the concepts relevant to an analysis.

*Identifiers.* Event tables (measurement, drug exposure, observation) use database-generated identifiers, because several source tables feed each of them. Patient and visit tables keep the source identifiers (MIMIC-III subject_id, hadm_id and icustay_id; eICU-CRD patientunitstayid, with person identifiers derived from uniquepid).

*Default concept IDs.* All concept ID columns default to 0 in Bronze, indicating unmapped concepts. The Silver layer assigns standard OMOP concept IDs from the curated source_to_concept_map. Records that remain at concept_id = 0 after Silver processing are excluded from the Gold analysis.

*Relaxed constraints.* Date columns are derived from the corresponding datetime columns and remain mandatory, except for the end date and datetime of visit_detail and the start and end dates of drug_exposure, which are missing in parts of the source data. Several text columns were widened to hold long source values, and vocabulary foreign keys were omitted because the full OMOP vocabulary is not loaded.

### S1.3 Layer Details

**Bronze Layer.** The Bronze layer maps source table structures into the OMOP CDM schema with one mapping step per source table. Each source table (e.g., MIMIC-III chartevents, eICU-CRD vitalperiodic) is handled by its own class in the code, which implements the column-level mapping. The set of mapping steps is fixed per source; JSON configuration files specify the database connection and row-level source filters, while the selection of parameters takes place in Silver.

For MIMIC-III, 13 mapping steps transfer the clinical source tables to seven OMOP target tables, and a further step fills the source_to_concept_map from the data dictionaries. For eICU-CRD, 16 source tables are mapped analogously, with admissiondrug used for drug-name lookup; offset-based timestamps are converted to absolute datetimes relative to a synthetic ICU admission time. All clinical concept identifiers (`measurement_concept_id`, `drug_concept_id`, `observation_concept_id`) are set to 0 in Bronze, deferring semantic interpretation to Silver. Original source identifiers, including MIMIC-III ITEMIDs (stored as text in `measurement_source_value`) and eICU-CRD parameter names, are kept in the corresponding `*_source_value` fields for traceability.

Quality assurance comprises automated Bronze tests (MIMIC-III and eICU-CRD) validating row counts, NULL constraints, foreign key integrity, source value preservation, timestamp consistency and schema extensions. Data quality filters are applied during extraction (e.g., exclusion of MIMIC-III chartevents flagged as erroneous by clinical staff).

*Bronze profiling.* After the Bronze ETL, an automated profiling step generates a statistical profile with one row per combination of source parameter and unit. If a parameter (e.g., $FiO_2$) appears with several units (e.g., "%" and "ratio"), each combination receives its own row with separate statistics. This prevents mixing distributions across incompatible units and provides the evidence base for unit standardization. A merge mode re-profiles the data while keeping existing annotations.

The profiler joins measurement statistics with parameter metadata from the `source_to_concept_map`, which Bronze fills from the source data dictionaries (MIMIC-III: `d_items`, `d_labitems`, `d_icd_diagnoses`, `prescriptions`). Where these dictionaries contain standardized terminology codes, Bronze keeps them as `target_concept_code`: LOINC codes from `d_labitems`, ICD-9 codes from `d_icd_diagnoses` and NDC codes from `prescriptions`. These codes carry through into the profiling CSV and give experts a starting point; parameters without standard codes, mainly bedside measurements from `chartevents`, are mapped manually. The profile is exported as a 45-column CSV with five column groups: (1) identification (source code, label, vocabulary, source table), (2) unit inventory (one row per observed unit), (3) mapping columns (target concept, target unit, conversion method, pre-filled where standardized codes exist), (4) annotation fields (include flag, plausibility range, row filter, analytical tags) and (5) descriptive statistics (value distributions, text values, temporal patterns, per-visit frequencies, storetime analysis).

This CSV serves as both a quality gate and the annotation template for Silver. The clinical parameters in the profile were identified in three steps: (1) compilation of all parameter identifiers from the six source code repositories analysed in [3] into a union set, supplemented for MIMIC-III by a search of its data dictionary structured by SOFA subscore; (2) mapping to standard OMOP concepts via the Athena vocabulary browser; and (3) validation by intensivists, who reviewed per-parameter statistics for clinical plausibility, correct SOFA subscore assignment and unit consistency. The validated union set comprises 349 source codes for MIMIC-III and 255 for eICU-CRD, covering the SOFA inputs together with ventilation, body weight, antibiotics, cultures and infection diagnoses. Because a single parameter may appear with multiple units, these correspond to 465 parameter-unit profiles for MIMIC-III and 641 for eICU-CRD. Standardized terminology codes (NDC, LOINC and ICD-9 in MIMIC-III; HICL and ICD-9 in eICU-CRD) were pre-populated for 49.5 % of the MIMIC-III and 10.6 % of the eICU-CRD profiles and only had to be verified; the standard OMOP concept was assigned to every profile in the annotation step. Repository-specific parameter sets were preserved for the D1 analysis.

Experts complete the mapping columns: OMOP concept IDs, target units, conversion methods (e.g., normalization of $FiO_2$ to percent), plausibility ranges, inclusion flags and analytical tags (e.g., `sepsis3_mimic`, `weight`, `infection`). The Silver ETL imports the annotated CSV as the authoritative `source_to_concept_map`, which makes every mapping traceable from raw source parameter to standard OMOP concept. The validated parameter list and plausibility ranges are provided in Supplement S2 (Section S2.2).

**Silver Layer.** The Silver layer copies the Bronze tables and then standardizes them with selective in-place SQL updates, so that all Bronze data are preserved and each step can be verified on its own. The steps run in this order: validation that every conversion method named in the annotated CSV is implemented; creation of the schema; full copy of the Bronze tables; constraints and indices; import of the annotated CSV, including tags; correction of unit labels; assignment of OMOP concept IDs; value standardization; removal of implausible values; assignment of unit concept IDs; and a summary of row counts and concept coverage. Value standardization covers $FiO_2$ normalization to percent, parsing of GCS subscores and of textual GCS entries, and the conversion of vasopressor infusion rates to $\mu$g/kg/min or units/min using body weight and infusion duration. Implausible measurement values are set to NULL in place. Deduplication takes place in Gold.

After the Silver ETL, a profiling step computes concept-level statistics on the standardized data (value distributions per OMOP concept, unit consistency, coverage) as a quality gate for concept mapping and unit standardization (results for the SOFA parameters in Supplement S2, Section S2.2).

**Gold Layer.** The Gold layer computes SOFA scores and Sepsis-3 labels from the standardized Silver data. The preprocessing decisions of domains D2–D5 and the parameter set (D1), which is chosen through the tags and concepts to load, are specified in one JSON configuration file per configuration.

*Cohort and data retrieval.* Cohort selection applies the configured filters (e.g., age, length of stay, first ICU stay, MetaVision documentation in MIMIC-III, and the exclusion criteria of the reconstructed repositories such as clinical services or treatment withdrawal) and stores the cohort in the OMOP `cohort` tables. A single query then combines four OMOP Silver tables into one long-format table (SQL UNION ALL). Each domain (measurement, drug_exposure, observation, death) is projected onto a common schema: `icustay_id` (from `visit_detail_id`), `concept_id`, `starttime`, `endtime`, `storetime`, `value`, `unit` and `value_str`. Tag-based filtering restricts retrieval to the concepts tagged for the current analysis. Age at admission is computed from `person.year_of_birth`, and ICU admission time from `visit_detail_start_datetime`.

*Cleaning.* The layer removes exact duplicate records, converts $FiO_2$ from percent, as stored in Silver, to fractions for the $PaO_2/FiO_2$ ratio and removes values outside harmonized plausibility ranges, so that the effective range is the narrower of the Silver and Gold limits. It derives ventilation episodes, sets the time origin (ICU admission in the reference) and, for stays with suspected infection, restricts the data to the window from 48 h before to 24 h after its onset.

*Vasopressor normalization.* Doses of seven vasopressors and inotropes (norepinephrine, epinephrine, dopamine, dobutamine, phenylephrine, vasopressin and milrinone) are expressed in $\mu$g/kg/min or units/min. Rows dosed in mg, which occur in CareVue and thus only when CareVue stays are included, are converted in Gold using the closest prior body weight of the ICU stay, with a default weight (80 kg in the reference) when no weight is recorded; all other rate units are converted in Silver.

*Temporal aggregation (D2/D3).* After pivoting to wide format (one column per clinical concept), measurements are aggregated into sliding windows anchored at ICU admission. In the reference, a 24 h window advances in 1 h steps; during the first 24 h, the window covers all data since admission. Aggregation functions are configurable per parameter. In the reference, each parameter is reduced to its most pathological value in the window, which is the maximum for bilirubin, creatinine, $FiO_2$ and vasopressor doses and the minimum for mean arterial pressure, $PaO_2$, platelets and GCS. $SpO_2$ is averaged, and urine output is summed.

*Imputation (D4).* A three-level strategy addresses missing values: (1) first-level derivation computes values from related measurements ($PaO_2$ from $SpO_2$ via a lookup table covering $SpO_2$ 80–100 %; GCS total from eye, verbal and motor subscores), (2) second-level forward filling carries the last observation forward, and (3) third-level defaults fill remaining gaps with constant default values, in the reference normal values (e.g., $FiO_2$ = 0.21 for room air, GCS = 15 for alert patients).

*SOFA computation (D5).* Six organ-specific subscores (respiratory, cardiovascular, hepatic, coagulation, renal, neurological) are computed per time window according to the SOFA criteria [4], and the total SOFA score is their sum. Configurable variants include $PaO_2/FiO_2$ versus $SpO_2/FiO_2$ (respiratory), dose-dependent versus binary vasopressor scoring (cardiovascular), with versus without urine output (renal) and the treatment of sedated or intubated patients (neurological). The baseline of the SOFA increase is configurable as the rolling minimum over a preceding window (24 h in the reference), the cumulative minimum since admission, the admission SOFA or zero; the increase can be restricted to measured subscores.

*Suspected infection (D6).* In MIMIC-III, infection is identified with the operationalization of Seymour et al. [5]: an antibiotic prescription (OMOP concept 4085730) combined with a microbiology culture (concept 4189544) obtained within 72 h before or 24 h after the antibiotic. In eICU-CRD, where microbiology culture data are too sparse for this criterion, infection is identified from sepsis diagnosis records (ICD-9/10 codes and the APACHE admission diagnosis, mapped to the OMOP concepts

44820254 and 44837445), as in the analysed eICU-CRD repositories. The route is selected per configuration (`antibiotic_culture`, `diagnosis` or `diagnosis_sofa`). For each route, the onset is computed both from documentation time and from measurement time.

*Sepsis-3 labelling.* A binary label is assigned when a SOFA increase of at least 2 points above the baseline falls within the window around suspected infection. On the `diagnosis` route of eICU-CRD, the diagnosis itself is the label and no SOFA criterion applies; `diagnosis_sofa` additionally requires the SOFA increase. Labels are computed for both readings of the infection onset.

*Profiling and export.* After the Gold pipeline, a profiling step computes per-column summary statistics (minimum, maximum, mean, median, quartiles, standard deviation, proportion missing) on the database server. A summarization step adds the per-stay prevalence of each label and the list of septic ICU stays with the hour of first onset. Each Gold result table holds one row per time window and ICU stay. Instead of transferring these tables, an export script combines the summaries of all configurations into CSV files of detection rates, SOFA subscore means, imputation fractions and septic stays. The analysis computes Jaccard indices from the septic-stay lists and differences in detection rates and mean subscores from the summary statistics.
