## Supplement S2: Configurations and Detailed Results for "Effect of Data Preprocessing Decisions on Sepsis-3 Labels in MIMIC-III and eICU-CRD: A Sensitivity Analysis"

This supplement provides the variant catalogue, the configurations and detailed results.

### S2.1 Preprocessing Variant Catalogue

The previous systematic review [1] identified 321 implementation decisions in the preprocessing domains D1–D6 (264 in MIMIC-III, 57 in eICU-CRD) across the four MIMIC-III and two eICU-CRD repositories. Its full decision catalogue additionally records 47 cohort-selection and 11 other decisions (379 in total). Many of these decisions are repository-specific instances of the same preprocessing method. For example, several repositories each restrict laboratory data to a time window around suspected infection, which is one D2 method applied several times. Others are implementation details that cannot be varied independently, such as specific ITEMID lists, which belong to the parameter selection (D1).

To derive testable variants, we consolidated the 321 decisions in three steps. Decisions that implement the same method in different repositories were merged into one variant. The D1 decisions were consolidated into repository-specific parameter sets (D1-S1 to D1-S6) instead of single ITEMID-level decisions. Implementation details that do not affect sepsis labels on their own, such as sort order, data type conversions and comments, were excluded. The 47 cohort-selection decisions are covered by a fixed cohort definition and are reproduced in the Tier 2 reconstructions. The 11 other decisions, such as severity-score comparisons and model architectures, lie outside the framework of D1–D6.

This consolidation yielded 51 variants in the five varied domains, namely 6 repository-specific parameter sets (D1), 10 temporal windows (D2), 3 aggregation rules (D3), 11 settings for missing values (D4) and 21 settings of the SOFA calculation (D5). Each variant was tested on its own against the reference configuration. Infection identification (D6) and the cohort definition were held fixed (Tables 6 and 7), apart from one stricter MIMIC-III infection rule (D6-S2) that is reported as a contrast in Section S2.4. All configurations use each measurement from its measurement time, as the reviewed repositories do. The contrast with documentation-time availability is reported in Section S2.4, together with the cohort and the time origin.

To avoid confusion with the decision identifiers of the previous review (D1-01, D2-01, ...), the variants of this paper are labelled D*d*-S*n*, for domain *d* and setting *n*. In D2–D5, setting 1 is the reference. In D1, the reference is the union set (D1-S0), and the repository sets are D1-S1 to D1-S6. Tables 1–7 list all variants, and ⋆ marks those that form the reference configuration. All listed variants were implemented and run.

The tables do not contain every method reported in the reviewed literature. Population-level imputers and missingness-based case exclusion (D4), most-recent-value aggregation (D3) and a modified SOFA (D5) were not tested, for reasons given in the respective domain sections. Measurements that the source system flagged as erroneous (`chartevents.error`, `outputevents.iserror`) are excluded in all configurations as a data-quality step and are therefore not listed as a variant.

**D1 — Parameter selection.** Each variant adopts the SOFA-relevant parameter set of one reviewed repository, that is, the measurements it extracts and assigns to each SOFA component. It is compared with the reference (D1-S0), the union of all validated parameters of the six repositories (Table 1). The MIMIC-III repositories cover between 125 and 143 ITEMIDs [1]. The repositories are sepsis3-mimic [2] (D1-S1), MIMIC-Code [3] (D1-S2), sepsis-3-labels [4] (D1-S3) and AI Clinician [5] (D1-S4) for MIMIC-III, and AI-enrichment [6] (D1-S5) and trajPLT [7] (D1-S6) for eICU-CRD.

**Table 1.** D1 — Parameter selection variants.

| ID | Variant | Description |
|---|---|---|
| D1-S0⋆ | Union set (reference) | Union of all validated SOFA parameters across the six repositories |
| D1-S1 | sepsis3-mimic | Repository-specific SOFA parameter set (MIMIC-III) |
| D1-S2 | MIMIC-Code | Repository-specific SOFA parameter set (MIMIC-III) |
| D1-S3 | sepsis-3-labels | Repository-specific SOFA parameter set (MIMIC-III) |
| D1-S4 | AI Clinician | Repository-specific SOFA parameter set (MIMIC-III) |
| D1-S5 | AI-enrichment | Repository-specific SOFA parameter set (eICU-CRD) |
| D1-S6 | trajPLT | Repository-specific SOFA parameter set (eICU-CRD) |

**D2 — Temporal windowing.** Two temporal settings are distinguished. The *aggregation window* is the span of data summarized into one SOFA evaluation, and the *observation period* is the part of the stay in which the label is evaluated. The reference (D2-S1) uses a 24-hour aggregation window that advances in 1-hour steps over the entire ICU stay, starting at the time origin $t_0$ = ICU admission. A SOFA increase counts only from 48 h before to 24 h after the onset of suspected infection (Methods). The time origin is not a D2 variant, and anchoring $t_0$ at the first measurement is reported in Section S2.4. The variants change the size of the sliding window (D2-S2 to D2-S5, all at a 1-hour step), replace it by non-overlapping blocks (D2-S6 and D2-S7) or change the placement of the evaluation, that is, the observation period, the anchor and the grid alignment (D2-S8 to D2-S11; Table 2). Window sizes of 1 to 24 h and the infection-centred anchor are taken from the repositories and the reviewed methods, and the 48 h window and the non-overlapping blocks extend them to the reported extremes. Reported window sizes range from about 30 min to 144 h. We sampled this range at 1, 4, 12, 24 and 48 h. The 30 min window lies below the 1-hour grid, and the longest reported windows (72 to 144 h) were not sampled.

The observation period is varied in both directions. The reference evaluates the whole stay, whereas D2-S8 (first 24 h only) reproduces the 24-hour horizon of four reviewed repositories (sepsis3-mimic, MIMIC-Code, AI-enrichment, trajPLT). The infection window of the reference follows sepsis3-mimic, AI Clinician and sepsis-3-labels. D2-S9 (no infection window) evaluates the SOFA increase over the whole stay, as the repositories without such a window do. D2-S10 (admission + infection) is an additional variant that is not taken from a single repository. It evaluates the stay from 4 h before to 24 h after ICU admission and keeps only stays whose first suspected infection lies between 48 h before and 24 h after admission (MIMIC-III 12,932 of 13,388 stays, eICU-CRD 93,619 of 94,782). It tests whether anchoring the label at admission and at infection at the same time changes the labels.

The aggregation offset is the step at which the sliding window advances and the SOFA score is re-evaluated. It is therefore the resolution at which a SOFA increase of at least 2 points can be detected, and it cannot be set to zero. The step is held at 1 h, the finest resolution reported. The coarser steps reported in the literature (3 to 24 h) are represented by the non-overlapping blocks (D2-S6, D2-S7). All D2 variants use measurement-time availability.

**Table 2.** D2 — Temporal windowing variants.

| ID | Variant | Description |
|---|---|---|
| *Window size (sliding)* | | |
| D2-S1⋆ | 24h (reference) | 24-hour sliding window, 1-hour step (offset), entire ICU stay; SOFA increase within −48h to +24h of suspected infection |
| D2-S2 | 1h | 1-hour sliding window, 1-hour step |
| D2-S3 | 4h | 4-hour sliding window, 1-hour step |
| D2-S4 | 12h | 12-hour sliding window, 1-hour step |
| D2-S5 | 48h | 48-hour sliding window, 1-hour step |
| *Non-overlapping blocks* | | |
| D2-S6 | 1h blocks | Non-overlapping 1-hour blocks |
| D2-S7 | 4h blocks | Non-overlapping 4-hour blocks |
| *Anchor, extent & grid alignment* | | |
| D2-S8 | First 24h only | Observation period limited to the first 24 hours after ICU admission (24-hour sliding window) |
| D2-S9 | No infection window | SOFA increase counted over the whole stay, not only within −48h to +24h of suspected infection |
| D2-S10 | Admission + infection | $-4$ to $+24$ h around admission; first suspected infection within $-48/+24$ h of admission (additional variant) |
| D2-S11 | 1h grid ceiling | Same anchor and 24h window as the reference, but the window-grid origin is snapped (ceiled) to a 1-hour wall-clock boundary |

**D3 — Aggregation strategy.** Within each window, the repeated measurements of a parameter are reduced to one value. The reference (D3-S1) takes the most pathological value of each parameter, which is the maximum for bilirubin, creatinine, $FiO_2$ and vasopressor dose and the minimum for platelets, mean arterial pressure, GCS and $PaO_2$. $SpO_2$ is averaged, and urine output is summed over 24 h. All six repositories make this choice. Two variants replace the worst value by a mean, as reported in the wider literature. D3-S2 averages the continuous parameters, which also covers the mean of laboratory values described in the review, and D3-S3 averages every parameter, vasopressors included. D3-S4 takes the least pathological value of each parameter. It is not a repository setting but a lower bound that, together with the reference, brackets the possible range. Outlier handling is the same in all D3 variants. Implausible values are removed before aggregation with clinician-validated plausibility ranges (Table 9), which is a data-quality step like the error-flag filtering and not a variant. Filtering after aggregation is supported by the pipeline but used by no reviewed repository. Median, first-value and most-recent aggregation were not evaluated. In the reviewed code, the median appears only for the imputation of comorbidities and the first value only in two eICU-CRD association studies as an extraction shortcut. A most-recent value would miss the transient organ dysfunction that the worst-value reference captures.

**Table 3.** D3 — Aggregation strategy variants.

| ID | Variant | Description |
|---|---|---|
| D3-S1⋆ | Worst-case (reference) | Parameter-specific most-pathological value (max bilirubin/creatinine/$FiO_2$/vasopressors, min platelets/MAP/GCS/$PaO_2$, mean $SpO_2$, 24-hour sum urine) |
| D3-S2 | Mean (continuous) | Mean of continuous parameters |
| D3-S3 | Mean incl. vasopressors | Uniform mean applied to every parameter, vasopressors included |
| D3-S4 | Best-case | Parameter-specific least-pathological value; labelled sanity-check lower bound |

**D4 — Missing-value handling.** Missing values are filled in three levels that are applied in order. Level 1 derives a value from related parameters (e.g., $PaO_2$ from $SpO_2$ via a lookup table, total GCS from its eye, verbal and motor subscores). Level 2 carries the last observation forward (LOCF). Level 3 fills any parameter that is still missing with a constant normal value, with the exception of urine output described below. The reference (D4-S1) applies all three levels, and the variants switch off or replace one level at a time (Table 4). For level 1, D4-S2 omits the derivation, and D4-S3 restricts the $PaO_2/FiO_2$ ratio to arterial blood gases with a specimen classifier. For level 2, D4-S4 omits forward filling, D4-S5 interpolates linearly, and D4-S6 limits LOCF to 6 h. For level 3, D4-S7 omits the default fill, D4-S8 fills with zero, D4-S9 uses near-normal constants, D4-S10 fills with the cohort mean, and D4-S11 treats unmeasured urine output as anuria. D4-S12 switches off all levels. The four variants that switch off levels (D4-S2, D4-S4, D4-S7, D4-S12) isolate the contribution of each level, and the remaining variants are taken from the reviewed repositories. The mean fill (D4-S10) is reported by five MIMIC studies. It fills each remaining gap with the cohort mean of the parameter, a value estimated from the whole dataset that a real-time system would not have. The constants of the reference instead encode the assumption that an unmeasured parameter indicates normal organ function. Median fill, reported by one study, is represented by D4-S10, because a single central value changes the SOFA inputs very little whether it is the mean or the median. D4-S9 moves the constants of the reference towards a typical but not perfectly healthy patient (e.g., creatinine 1.2 and bilirubin 1.0 mg/dL).

The default for urine output needs separate treatment. The renal subscore reaches its two highest grades through either creatinine or oliguria, so the value that fills an unmeasured urine output decides this subscore for every stay in which urine was never charted. In practice, these are stays without an indwelling urinary catheter, which are not the most severely ill. The repositories differ here. Four of the six (sepsis3-mimic, MIMIC-Code, sepsis-3-labels, AI Clinician) set no default, so that the renal subscore relies on creatinine, whereas the two eICU-CRD repositories (AI-enrichment, trajPLT) fill with zero, which scores an unmeasured output as anuria. The reference follows the majority and treats an unmeasured urine output as unknown. This is consistent with the assumption behind the other level-3 constants, because a zero volume would be the only constant that asserts maximal organ dysfunction. Filling with zero is kept as a separate variant (D4-S11) that changes the urine default alone, unlike D4-S8, which fills every level-3 gap with zero.

KNN, MICE and predictive mean matching (PMM) fill the gaps of a stay from other stays, either from a pool of donors (KNN) or from a model fitted across the dataset (MICE, PMM). The pipeline instead imputes within each stay. Both possible implementations are unsuitable for this study. A leave-one-out donor pool scales quadratically, cannot be computed stay by stay and presumes a pool that a real-time system does not have. A held-out donor subset would have to be excluded from every configuration, because all settings are compared on the same cohort, which would cost a large share of the stays. These methods do occur in the reviewed literature (KNN in AI Clinician [5], multiple imputation in five studies, PMM in four).

For the same reason, we did not exclude stays whose missingness exceeds a threshold, as several studies do at thresholds of 10 to 50 %. Removing stays would change the cohort between configurations and mix preprocessing effects with cohort composition. Such exclusions and minimum-measurement requirements are applied only in the Tier 2 reconstructions, where they belong to the reconstructed pipeline (e.g., `exclude_missing_data` and the minimum number of urine measurements in sepsis3-mimic).

**Table 4.** D4 — Missing-value handling variants.

| ID | Variant | Description |
|---|---|---|
| D4-S1⋆ | Three-level (reference) | Derivation + LOCF + constant-normal default fill |
| *Level 1 — derivation* | | |
| D4-S2 | No derivation | Skip $PaO_2$-from-$SpO_2$ and GCS-from-subscores derivation |
| D4-S3 | Arterial-specimen classifier | Restrict the $PaO_2/FiO_2$ ratio to arterial blood-gas specimens (only arterial $PaO_2$ feeds the ratio) |
| *Level 2 — primary imputation* | | |
| D4-S4 | No forward fill | Disable LOCF |
| D4-S5 | Linear interpolation | Interpolate between observations instead of LOCF |
| D4-S6 | Time-limited 6h | LOCF limited to a 6-hour carry-forward horizon |
| *Level 3 — default fill* | | |
| D4-S7 | No constants | Disable constant-normal default fill |
| D4-S8 | Zero fill | Fill remaining gaps with zero instead of normal values |
| D4-S9 | Near-normal constants | Level-3 constants of a typical rather than perfectly healthy patient (creatinine 1.2, bilirubin 1.0 mg/dL); alternative constants |
| D4-S10 | Mean fill | Fill remaining Level-3 gaps with the parameter's cohort mean instead of a clinical-normal constant |
| D4-S11 | Unmeasured urine as anuria | Fill an unmeasured urine output with zero, scoring it as anuria (eICU convention); the reference leaves it unknown so the renal subscore falls back on creatinine |
| *All levels off* | | |
| D4-S12 | No imputation | All three levels disabled |

**D5 — SOFA calculation.** The reference (D5-S1) combines the standard SOFA choices [8], namely ventilation-dependent respiratory thresholds, dose-dependent cardiovascular scoring, a renal score from creatinine and urine output, the standard GCS and a rolling 24-hour minimum as baseline of the SOFA increase. The variants change the scoring of one organ system or the baseline and onset definition (Table 5). The respiratory variants are a ventilation-independent $PaO_2/FiO_2$ score (D5-S2) and a direct $SpO_2/FiO_2$ substitution (D5-S3). The cardiovascular variants are binary scoring of any vasopressor (D5-S4), norepinephrine-equivalent dosing (D5-S5) and the limited vasopressor set of trajPLT (D5-S6). The CNS variants are a GCS of 15 for sedated patients (D5-S7), a verbal score of 0 for intubated patients (D5-S8) and the worst GCS within 4 h (D5-S9). The renal variants are renal replacement therapy as maximal dysfunction (D5-S10), 4-hour urine-output thresholds (D5-S11) and creatinine only (D5-S12). The baseline and onset variants are the cumulative minimum since admission (D5-S13), the admission value (D5-S14), a zero baseline (D5-S15), first onset only (D5-S16), early-onset exclusion (D5-S17) and baseline windows of 1, 2, 4, 6 and 12 h (D5-S18 to D5-S22).

The baseline is the one reference setting that is not taken from the reviewed repositories, which use a zero, admission or cumulative-minimum baseline. Only 2 of the 15 MIMIC-III studies that described their SOFA calculation documented a baseline at all [1]. We chose the rolling 24-hour minimum because it reads Sepsis-3 as an acute deterioration that can be detected at any hour of the stay. It corresponds to the "two-point deterioration in SOFA score within a 24-hour period" of the PhysioNet/Computing in Cardiology Challenge 2019 [9] and is available as an option in ricu [10].

All variants are taken from the reviewed repositories or reported methods. D5-S2, D5-S3, D5-S6 and D5-S12 follow trajPLT [7], with the $SpO_2/FiO_2$ bands of D5-S3 taken from Jones et al. [11]. Other equivalences between $SpO_2/FiO_2$ and $PaO_2/FiO_2$ exist, notably the SOFA-specific thresholds of Pandharipande et al. [12] and the linear mapping of Rice et al. [13]. They can shift the respiratory subscore and were not varied further. D5-S13 follows sepsis-3-labels [4]. Excluding dobutamine from cardiovascular scoring, as in sepsis3-mimic, changed detection rates by less than 0.1 pp and is not listed. The early-onset exclusion (D5-S17) removes sepsis that appears within the first hours after admission. It presumes that such onsets can be attributed to deterioration before the ICU stay, which is only possible in retrospect, so a real-time system could not apply it. A modified SOFA that changes two organ-scoring rules at once was not evaluated, because its effect could not be attributed to one decision.

The baseline definitions lie on one axis of strictness, because the Sepsis-3 criterion is a change and every implementation has to decide relative to what. A minimum over a longer span is lower and therefore easier to exceed by two points. A zero baseline requires no deterioration at all, an admission or cumulative-minimum baseline (D5-S14, D5-S13) measures deterioration since the beginning of the stay, and a trailing-window minimum measures only recent deterioration, with shorter windows requiring a faster rise. The window length is therefore a setting of its own and was sampled at 1, 2, 4, 6 and 12 h (D5-S18 to D5-S22). Six hours is the only change window reported in the reviewed literature [14], and

**Table 5.** D5 — SOFA calculation variants.

| ID | Variant | Description |
|---|---|---|
| D5-S1⋆ | Standard (reference) | Ventilation-dependent $PaO_2/FiO_2$, dose-dependent cardiovascular, creatinine + urine renal, standard GCS, rolling 24-hour minimum baseline |
| *Respiratory* | | |
| D5-S2 | Ventilation-independent $PaO_2/FiO_2$ | Assign the severe respiratory scores (SOFA 3–4) from the $PaO_2/FiO_2$ ratio alone, dropping the reference's mechanical-ventilation requirement for those scores |
| D5-S3 | $SpO_2/FiO_2$ substitution | Score the respiratory subscore directly from the $SpO_2/FiO_2$ ratio when $PaO_2$ is unavailable (distinct from the reference, which converts $SpO_2$ to a $PaO_2$-equivalent), using the $SpO_2/FiO_2$ bands of Jones et al. [11] |
| *Cardiovascular* | | |
| D5-S4 | Binary vasopressor | Any vasopressor = 3 points |
| D5-S5 | Norepinephrine-equivalent | Vasopressor doses converted to norepinephrine equivalents |
| D5-S6 | Limited vasopressor set | Cardiovascular score from dopamine, dobutamine, norepinephrine and epinephrine only (trajPLT) |
| *CNS* | | |
| D5-S7 | Sedated GCS = 15 | Assume GCS 15 for sedated patients |
| D5-S8 | Intubated verbal = 0 | Verbal score set to 0 for intubated patients |
| D5-S9 | Worst GCS in 4h | CNS score from the minimum GCS within a 4-hour lookback window |
| *Renal* | | |
| D5-S10 | CRRT as maximal dysfunction | Renal replacement therapy scored as 4 |
| D5-S11 | 4h urine thresholds | Urine-output thresholds evaluated over 4-hour windows |
| D5-S12 | Creatinine-only | Renal subscore from creatinine alone, without the urine-output criterion |
| *Sepsis-3 baseline & onset* | | |
| D5-S13 | Cumulative-minimum baseline | Baseline is the lowest SOFA since ICU admission |
| D5-S14 | Admission baseline | Increase ≥2 above the admission SOFA |
| D5-S15 | Zero baseline | SOFA ≥2 absolute (baseline = 0) |
| D5-S16 | First onset only | Count only the first sepsis episode per stay |
| D5-S17 | Early-onset exclusion | Exclude sepsis onset within the first 7h of ICU admission |
| D5-S18 | 1h baseline window | Baseline is the preceding hour, so only an abrupt step qualifies |
| D5-S19 | 2h baseline window | Baseline is the minimum of the preceding 2 hours |
| D5-S20 | 4h baseline window | Baseline is the minimum of the preceding 4 hours |
| D5-S21 | 6h baseline window | The ≥2-point rise must occur within 6 hours instead of the reference's 24 hours |
| D5-S22 | 12h baseline window | Baseline is the minimum of the preceding 12 hours |

a 1-hour window approximates a comparison from hour to hour. With a trailing window, the baseline rises together with a deteriorating patient, so a slow decline may never produce a two-point rise while a short fluctuation does, and this effect grows as the window shortens. The admission and cumulative-minimum baselines do not have this property. The Sepsis-3 consensus specifies the threshold and assumes a baseline of zero in the absence of known organ dysfunction [15] but prescribes no window. The baseline is also rarely reported. Of the 44 MIMIC-III studies in the previous review [1], two state a baseline method and one states a change window.

**D6 — Infection identification.** Suspected infection is not varied in Tier 1 and is held fixed within each database (D6-S1). One stricter MIMIC-III rule is reported as a contrast in Section S2.4. D6-S2 (four-day antibiotic course) counts a culture only if an antibiotic course of at least four days starts within two days of the culture day or lasts until the day before death, adapted from the CDC Adult Sepsis Event. Everything else, including the infection window, follows the reference. eICU-CRD has no such variant, because its reference takes infection from diagnosis records, to which the course rule does not apply.

The operationalization depends on the database, because MIMIC-III and eICU-CRD differ in which infection-related data are reliably recorded. In MIMIC-III, suspected infection follows Seymour et al. [16], that is, an antibiotic combined with a culture obtained within 72 h before or 24 h after it. The antibiotic and culture lists of the repositories were not copied but passed through the same profiling and annotation workflow as every clinical parameter (Supplement S1, Section S1.3). Each antibiotic and microbiology source concept used by an analysed repository is tagged by repository in the `source_to_concept_map`, and clinical experts reviewed every tagged concept against its profile (units, dose quantities, value distributions and administration frequencies) before inclusion. Because microbiology documentation is severely underrepresented in the older CareVue system [17], culture-based infection detection is unreliable for CareVue stays, and the MIMIC-III analysis is restricted to MetaVision stays (cohort criterion C4). This restriction follows sepsis3-mimic. According to the source-code analysis of the previous review [1], that pipeline identifies 25 suspected infections among CareVue stays and 18,423 among MetaVision stays and consequently excludes CareVue (37,912 stays, 61.6 % of its cohort).

In eICU-CRD, this criterion cannot be applied. Microbiology cultures are sparsely recorded, so the Seymour operationalization yields a suspected-infection rate near zero that reflects missing culture data. Suspected infection is therefore identified from sepsis diagnosis records, namely ICD-9/10 codes and the APACHE admission diagnosis (`apacheadmissiondx`), mapped to the OMOP concepts 44820254 and 44837445, and combined with the same SOFA criterion. The two analysed eICU-CRD repositories take infection from these sources. trajPLT uses ICD codes or the APACHE admission diagnosis, and AI-enrichment uses the APACHE admission diagnosis alone. Because the two sources are combined with a logical or, the APACHE admission diagnosis widens the set of infected stays and does not restrict labelling to infections coded at admission. As a result, infection is defined from diagnosis records in eICU-CRD and from the antibiotic and culture criterion in MIMIC-III, so absolute detection rates differ between the databases for this reason as well as because of the populations.

Each database is thus handled with the infection definition of its repositories. MIMIC-III has a subpopulation with valid culture data (MetaVision), so the Seymour operationalization is kept and the cohort restricted. In eICU-CRD, cultures are sparse throughout, so no cohort restriction can recover the criterion. ICD-based infection is not an established Sepsis-3 operationalization for MIMIC-III. None of the four analysed MIMIC-III repositories uses it, and only two of the reviewed MIMIC-III studies report it, so we did not apply it to CareVue stays. To quantify the effect of the MetaVision restriction, we ran the reference configuration on CareVue stays and on all MIMIC-III stays with the same culture-based definition (Section S2.4). The detection rate was 31.5 % in MetaVision, 22.4 % in CareVue and 26.2 % in both together, which is consistent with the underrepresented microbiology in CareVue.

Documentation-time availability is not a D6 variant, because it concerns when a measurement becomes available and not how infection is defined. It is reported in Section S2.4.

**Table 6.** D6 — Infection identification. Held fixed per database (D6-S1; not varied in Tier 1); the operationalization differs by database because culture-based infection markers are reliably populated only in the MIMIC-III MetaVision subpopulation. D6-S2 is a single MIMIC-III contrast (Section S2.4).

| ID / Database | Operationalization | Description |
|---|---|---|
| D6-S1⋆ MIMIC-III | Seymour et al. [16] | Co-occurring antibiotic administration and microbiology sampling (culture 72 h before to 24 h after antibiotics); clinically validated antibiotic and culture criteria; MetaVision stays only (CareVue microbiology underrepresented [17]) |
| D6-S1⋆ eICU-CRD | Sepsis diagnosis records | Infection from sepsis ICD-9/10 codes or the APACHE admission diagnosis (OMOP concepts 44820254 and 44837445), plus the SOFA criterion; culture-based Seymour not viable (culture data too sparse → near-zero detection) |
| D6-S2 MIMIC-III | Four-day antibiotic course | As D6-S1, but a culture counts only with an antibiotic course of ≥4 days starting within 2 days of it, or lasting until the day before death (adapted from the CDC Adult Sepsis Event) |

**Cohort definition.** All configurations share one reference cohort, so that differences in detection rate reflect preprocessing and not cohort composition. The reference includes adults (≥18 years, C1), the first ICU stay of each patient (C2) and stays longer than 24 h (C3), and for MIMIC-III only MetaVision stays (C4, see D6). These criteria are common to the analysed repositories and are held fixed (Table 7). Three further cohort operations of individual repositories were not adopted. Random subsampling serves computational tractability in some pipelines and does not change the expected detection rate. Disease-specific exclusions, such as the exclusion of cardiac surgery in sepsis3-mimic (5,905 stays, 9.6 %), are specific to a study, and we kept a general ICU cohort. Early-onset exclusion is included as a variant of sepsis onset (D5-S17) and not as a cohort filter, because it changes which onsets are counted and not which patients are eligible. The Tier 2 reconstructions use the cohort definition of each repository, including its exclusions. Their cohort sizes and absolute detection rates therefore differ between repositories and are not directly comparable with the fixed-cohort rates of Tier 1.

**Table 7.** Reference cohort definition (held fixed across all configurations; not varied in Tier 1).

| ID | Variant | Description |
|---|---|---|
| C1⋆ | Age ≥ 18 | Minimum age at ICU admission |
| C2⋆ | First ICU stay only | Only first ICU stay per patient |
| C3⋆ | LOS > 24h | Minimum ICU length of stay |
| C4⋆ | MetaVision only | Exclude CareVue ICU stays. Microbiology cultures are severely underrepresented in the CareVue system [17], making reliable infection identification infeasible for CareVue stays. Default for MIMIC-III sepsis labelling. |

Legend: all variants listed above were implemented and run. ⋆ = reference configuration (Tier 1 baseline).

### S2.2 Data Harmonization and Plausibility Ranges

Table 8 summarizes what Bronze and Silver profiling (Supplement S1, Section S1.3) show for the SOFA parameters. The statistics cover all ICU stays of each database rather than the study cohort. The Silver profile pools all source codes of a concept, and values recorded at the same time under several codes of one concept, such as a laboratory value that is also charted at the bedside, count once; the intervals therefore describe the data as the Gold layer receives them. The table is generated by the analysis repository (`src/tables/tab_s2_harmonization.py`) from the Bronze and Silver profiling files, which will be released with it (`data/profiling`) together with the annotated mapping files upon publication.

**Table 8.** Harmonization and measurement density of the SOFA parameters, from the Bronze and Silver profiles over all ICU stays of each database. Codes: source codes mapped to the OMOP concept. Units: distinct spellings of the source unit (e.g., K/uL, K/mcL, th/mm3 for platelets), all mapped to one target unit. Conv.: conversion methods applied in Silver (e.g., ml/h to $\mu$g/kg/min). Interval: mean time between consecutive values of the concept within an ICU stay, after pooling all source codes. Delay: mean time from measurement to documentation (MIMIC-III only; eICU-CRD records no documentation time).

| | MIMIC-III | | | | | eICU-CRD | | | |
|---|---|---|---|---|---|---|---|---|---|
| **Parameter** | Codes | Units | Conv. | Interval (h) | Delay (h) | Codes | Units | Conv. | Interval (h) |
| *Respiration* | | | | | | | | | |
| $PaO_2$ | 2 | 3 | 0 | 7.7 | 0.2 | 2 | 10 | 0 | 15.4 |
| $FiO_2$ | 1 | 1 | 1 | 17.9 | – | 4 | 5 | 1 | 2.5 |
| $SpO_2$ | 5 | 1 | 0 | 0.9 | 0.5 | 2 | 0 | 0 | 0.1 |
| *Coagulation* | | | | | | | | | |
| Platelets | 3 | 1 | 0 | 13.6 | 0.9 | 1 | 55 | 0 | 23.4 |
| *Liver* | | | | | | | | | |
| Bilirubin | 4 | 1 | 0 | 23.9 | 2.1 | 1 | 3 | 0 | 34.5 |
| *Cardiovascular* | | | | | | | | | |
| Mean arterial pressure | 7 | 1 | 0 | 0.9 | 0.5 | 2 | 0 | 0 | 0.2 |
| Norepinephrine | 2 | 3 | 1 | 1.5 | 0.5 | 19 | 5 | 3 | 4.0 |
| Epinephrine | 1 | 2 | 1 | 0.9 | 0.2 | 9 | 8 | 5 | 6.6 |
| Dopamine | 2 | 3 | 1 | 1.5 | 0.5 | 6 | 3 | 2 | 4.5 |
| Dobutamine | 2 | 3 | 1 | 1.3 | 0.4 | 7 | 6 | 2 | 5.9 |
| Phenylephrine | 2 | 3 | 1 | 1.4 | 0.4 | 11 | 7 | 4 | 3.9 |
| Vasopressin | 1 | 2 | 1 | 16.4 | 5.5 | 6 | 6 | 3 | 1.7 |
| Milrinone | 2 | 3 | 1 | 1.1 | 0.5 | 5 | 8 | 2 | 4.5 |
| *Central nervous system* | | | | | | | | | |
| GCS total | 1 | 1 | 1 | 3.3 | 0.6 | 2 | 0 | 1 | 4.1 |
| GCS eye | 2 | 0 | 2 | 3.4 | 0.7 | 5 | 0 | 1 | 8.5 |
| GCS verbal | 2 | 0 | 2 | 3.4 | 0.7 | 6 | 0 | 1 | 8.5 |
| GCS motor | 2 | 0 | 2 | 3.4 | 0.7 | 7 | 0 | 1 | 8.5 |
| *Renal* | | | | | | | | | |
| Creatinine | 4 | 1 | 0 | 12.0 | 1.4 | 1 | 3 | 0 | 36.7 |
| Urine output | 12 | 2 | 0 | 1.8 | 0.5 | 13 | 0 | 0 | 3.8 |
| *Other* | | | | | | | | | |
| Body weight | 5 | 1 | 0 | 26.5 | 0.7 | 1 | 0 | 0 | 26.5 |

The reference configuration removes physiologically implausible measurements before SOFA scoring (Table 9). The bounds were adopted from MIMIC-Extract [18], which takes its ranges from the mimic3-benchmarks suite [19], and each bound was reviewed for plausibility with an experienced anaesthesiologist. Ranges for catecholamines and vasoactive drugs, for which neither source specifies bounds, were defined from clinical dosing experience. The same ranges are used in the reference and in all Tier 1 and Tier 3 variants of both databases, so that differences between the databases cannot result from different cleaning. The Tier 2 reconstructions keep the ranges documented in each repository. Values outside the range are removed before aggregation, and drug rates are checked after normalization to weight-based units.

**Table 9.** Clinician-validated outlier plausibility ranges per SOFA-relevant parameter, applied in the reference and in all Tier 1 and Tier 3 variants of both databases.

| Parameter | OMOP concept ID | Unit | Lower | Upper |
|---|---|---|---|---|
| *Laboratory and vital signs* | | | | |
| Bilirubin (total) | 3024128 | mg/dL | 0.10 | 60.00 |
| Creatinine | 3016723 | mg/dL | 0.10 | 60.00 |
| Platelets | 3007461 | $10^3/\mu$L | 0.00 | 2000.00 |
| Mean arterial pressure | 37168599 | mmHg | 14.00 | 330.00 |
| $PaO_2$ | 3027801 | mmHg | 32.00 | 700.00 |
| $FiO_2$ | 42869590 | fraction | 0.21 | 1.00 |
| $SpO_2$ | 4020553 | % | 0.00 | 100.00 |
| *Catecholamines and vasoactive drugs* | | | | |
| Dopamine | 1337860 | $\mu$g/kg/min | 0.01 | 50.00 |
| Dobutamine | 1337720 | $\mu$g/kg/min | 0.01 | 50.00 |
| Epinephrine | 1343916 | $\mu$g/kg/min | 0.01 | 2.00 |
| Norepinephrine | 1321341 | $\mu$g/kg/min | 0.01 | 2.00 |
| Phenylephrine | 1135766 | $\mu$g/kg/min | 0.01 | 50.00 |
| Vasopressin | 1507835 | units/min | 0.01 | 10.00 |
| Milrinone | 1368671 | $\mu$g/kg/min | 0.01 | 50.00 |
| *Neurological score* | | | | |
| Glasgow Coma Scale (total) | 3007194 | points | 3 | 15 |

Concept identifiers are OHDSI OMOP standard concepts (Athena vocabularies). Glasgow Coma Scale component subscales use their definitional ranges (eye 1–4, verbal 1–5, motor 1–6). The Tier 2 reconstructions use the ranges of each repository, which are not listed here.

The full per-parameter source mapping (MIMIC-III ITEMIDs and eICU identifiers per OMOP concept, with units) will be released with the code upon publication; consistent with Section S2.1, ITEMID-level detail is not enumerated here, as it is subsumed by the D1 parameter-selection variants.

### S2.3 Detailed Results

The reference configuration combines all ⋆-marked variants from Section S2.1. Tier 1 variants vary one domain at a time while holding all others at this reference.

**All Tier 1 variants.** Figure 1 shows the detection rates of all 51 variants of D1–D5.

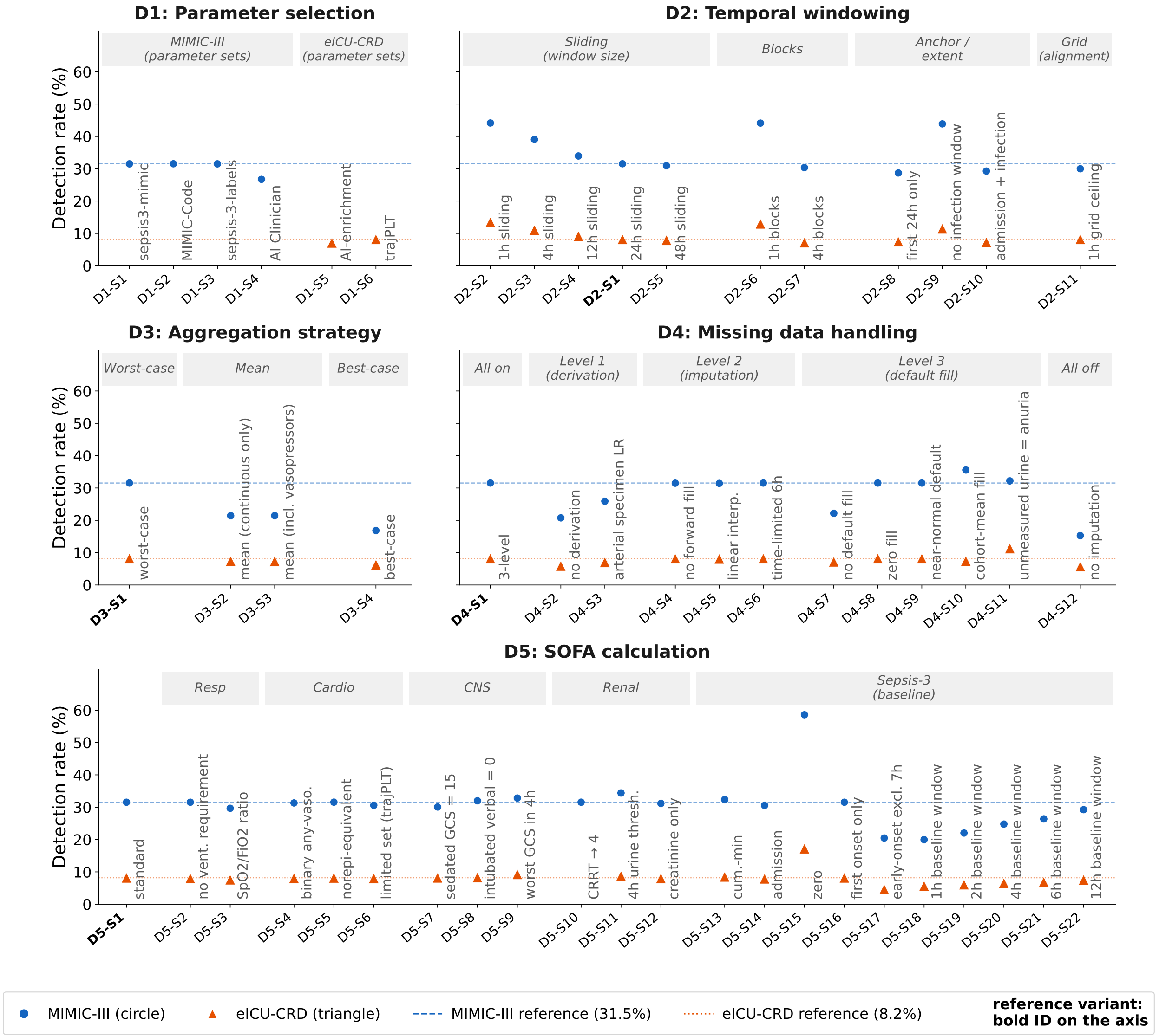


**Figure 1.** Sepsis-3 detection rates of all Tier 1 variants. In each panel (D1–D5), each data point is one variant, labelled by its identifier D$d$-S$n$ (domain $d$, setting $n$; Section S2.1); the identifier of each domain's reference variant is set in bold, and the grey headers group the variants as in the catalogue tables. All configurations share one cohort per database, $t_0$ = ICU admission and measurement-time availability. MIMIC-III circles, eICU-CRD triangles; dashed and dotted lines mark the reference rates.

Table 10 lists the detection rate, the difference from the reference and the Jaccard index of every variant of the processing domains in both databases; the main text shows the variants with the largest effects on rate or patients in a separate figure.

**Table 10.** All Tier 1 variants of the processing domains D2–D5. Rate in %. Δ is the difference from the reference in percentage points, + and − are the ICU stays that gain and lose the label compared with the reference, and $J$ is the Jaccard index of the septic stays against the reference (1 for the same stays, 0 for no common stays). The range of each domain is given in brackets (MIMIC-III / eICU-CRD), and a dash marks a variant that was not run in the database. [a]Listed but not counted in the range of its domain (lower bound or additional variant). The D1 parameter sets are given in Table 11.

| | | **MIMIC-III** | | | | | **eICU-CRD** | | | | |
|---|---|---|---|---|---|---|---|---|---|---|---|
| **ID** | **Variant** | **Rate** | **Δ** | **+** | **−** | ***J*** | **Rate** | **Δ** | **+** | **−** | ***J*** |
| *D2 Temporal windows (range 15.4 / 6.3 pp)* | | | | | | | | | | | |
| D2-S1 | 24h sliding (reference) | 31.5 | | | | | 8.2 | | | | |
| D2-S2 | 1h sliding | 44.1 | +12.6 | 1,828 | 140 | 0.67 | 13.5 | +5.3 | 5,380 | 348 | 0.56 |
| D2-S3 | 4h sliding | 39.0 | +7.5 | 1,125 | 121 | 0.77 | 11.1 | +2.9 | 3,191 | 457 | 0.67 |
| D2-S4 | 12h sliding | 34.0 | +2.4 | 375 | 53 | 0.91 | 9.2 | +1.0 | 1,105 | 176 | 0.86 |
| D2-S5 | 48h sliding | 30.9 | −0.6 | 8 | 89 | 0.98 | 7.9 | −0.2 | 49 | 282 | 0.96 |
| D2-S6 | 1h blocks | 44.1 | +12.6 | 1,826 | 142 | 0.67 | 13.0 | +4.8 | 4,988 | 427 | 0.57 |
| D2-S7 | 4h blocks | 30.4 | −1.2 | 748 | 906 | 0.67 | 7.2 | −1.0 | 1,560 | 2,514 | 0.56 |
| D2-S8 | first 24h only | 28.7 | −2.8 | 0 | 435 | 0.90 | 7.5 | −0.7 | 0 | 703 | 0.91 |
| D2-S9 | no infection window | 43.9 | +12.3 | 1,654 | 3 | 0.72 | 11.5 | +3.3 | 3,127 | 26 | 0.71 |
| D2-S10 | admission + infection[a] | 29.3 | −2.3 | 0 | 437 | 0.90 | 7.3 | −0.9 | 144 | 1,061 | 0.85 |
| D2-S11 | 1h grid ceiling | 30.0 | −1.5 | 109 | 315 | 0.90 | 8.2 | 0.0 | 0 | 0 | 1.00 |
| *D3 Aggregation (range 10.1 / 0.8 pp)* | | | | | | | | | | | |
| D3-S1 | worst-case (reference) | 31.5 | | | | | 8.2 | | | | |
| D3-S2 | mean (continuous only) | 21.4 | −10.1 | 84 | 1,437 | 0.65 | 7.4 | −0.8 | 561 | 1,307 | 0.78 |
| D3-S3 | mean (incl. vasopressors) | 21.4 | −10.1 | 91 | 1,442 | 0.64 | 7.4 | −0.8 | 567 | 1,331 | 0.77 |
| D3-S4 | best-case[a] | 16.8 | −14.7 | 39 | 2,007 | 0.52 | 6.3 | −1.9 | 737 | 2,513 | 0.62 |
| *D4 Missing values (range 20.3 / 5.6 pp)* | | | | | | | | | | | |
| D4-S1 | 3-level (reference) | 31.5 | | | | | 8.2 | | | | |
| D4-S2 | no derivation | 20.8 | −10.8 | 3 | 1,446 | 0.66 | 5.9 | −2.3 | 1 | 2,158 | 0.72 |
| D4-S3 | arterial specimen LR | 25.9 | −5.6 | 1 | 755 | 0.82 | 7.1 | −1.1 | 1 | 1,053 | 0.86 |
| D4-S4 | no forward fill | 31.5 | −0.1 | 9 | 16 | 0.99 | 8.2 | 0.0 | 91 | 110 | 0.97 |
| D4-S5 | linear interp. | 31.4 | −0.1 | 1 | 16 | 1.00 | 8.1 | −0.1 | 23 | 107 | 0.98 |
| D4-S6 | time-limited 6h | 31.5 | 0.0 | 0 | 0 | 1.00 | 8.2 | 0.0 | 0 | 0 | 1.00 |
| D4-S7 | no default fill | 22.1 | −9.4 | 0 | 1,259 | 0.70 | 7.2 | −1.0 | 0 | 936 | 0.88 |
| D4-S8 | zero fill | 31.5 | 0.0 | 0 | 0 | 1.00 | 8.2 | 0.0 | 0 | 0 | 1.00 |
| D4-S9 | near-normal default | 31.5 | 0.0 | 0 | 0 | 1.00 | 8.2 | 0.0 | 0 | 0 | 1.00 |
| D4-S10 | cohort-mean fill | 35.6 | +4.0 | 1,294 | 755 | 0.63 | 7.4 | −0.8 | 135 | 853 | 0.87 |
| D4-S11 | unmeasured urine = anuria | 32.2 | +0.7 | 87 | 1 | 0.98 | 11.3 | +3.1 | 2,979 | 31 | 0.72 |
| D4-S12 | no imputation | 15.3 | −16.3 | 4 | 2,183 | 0.48 | 5.7 | −2.5 | 3 | 2,329 | 0.70 |
| *D5 SOFA calculation (range 38.6 / 12.6 pp)* | | | | | | | | | | | |
| D5-S1 | standard (reference) | 31.5 | | | | | 8.2 | | | | |
| D5-S2 | no vent. requirement | 31.5 | 0.0 | 21 | 22 | 0.99 | 8.0 | −0.2 | 256 | 433 | 0.91 |
| D5-S3 | SpO2/FiO2 ratio | 29.7 | −1.9 | 73 | 325 | 0.91 | 7.6 | −0.6 | 117 | 666 | 0.90 |
| D5-S4 | binary any-vaso. | 31.3 | −0.2 | 0 | 30 | 0.99 | 8.1 | −0.1 | 24 | 131 | 0.98 |
| D5-S5 | norepi-equivalent | 31.6 | 0.0 | 10 | 8 | 1.00 | 8.2 | 0.0 | 21 | 41 | 0.99 |
| D5-S6 | limited set (trajPLT) | 30.6 | −1.0 | 145 | 273 | 0.90 | 8.1 | −0.1 | 63 | 181 | 0.97 |
| D5-S7 | sedated GCS = 15 | 30.1 | −1.5 | 40 | 237 | 0.94 | 8.2 | 0.0 | 124 | 115 | 0.97 |
| D5-S8 | intubated verbal = 0 | 32.0 | +0.4 | 124 | 64 | 0.96 | 8.3 | +0.1 | 197 | 84 | 0.96 |
| D5-S9 | worst GCS in 4h | 32.8 | +1.3 | 234 | 62 | 0.93 | 9.2 | +1.1 | 1,037 | 20 | 0.88 |
| D5-S10 | CRRT → 4 | 31.6 | 0.0 | 2 | 1 | 1.00 | – | – | – | – | – |
| D5-S11 | 4h urine thresh. | 34.4 | +2.9 | 390 | 7 | 0.91 | 8.7 | +0.6 | 584 | 56 | 0.92 |
| D5-S12 | creatinine only | 31.2 | −0.4 | 0 | 48 | 0.99 | 8.0 | −0.2 | 0 | 162 | 0.98 |
| D5-S13 | cumulative-minimum baseline | 32.4 | +0.8 | 109 | 0 | 0.97 | 8.5 | +0.3 | 296 | 0 | 0.96 |
| D5-S14 | baseline at admission | 30.6 | −1.0 | 80 | 211 | 0.93 | 7.9 | −0.3 | 240 | 509 | 0.91 |
| D5-S15 | zero baseline | 58.6 | +27.1 | 3,622 | 0 | 0.54 | 17.2 | +9.0 | 8,540 | 0 | 0.48 |
| D5-S16 | first onset only | 31.5 | 0.0 | 0 | 0 | 1.00 | 8.2 | 0.0 | 0 | 0 | 1.00 |
| D5-S17 | early-onset excl. 7h | 20.5 | −11.1 | 0 | 1,483 | 0.65 | 4.6 | −3.5 | 0 | 3,360 | 0.57 |
| D5-S18 | 1h baseline window | 20.0 | −11.6 | 0 | 1,547 | 0.63 | 5.7 | −2.5 | 0 | 2,393 | 0.69 |
| D5-S19 | 2h baseline window | 22.0 | −9.5 | 0 | 1,273 | 0.70 | 6.1 | −2.1 | 0 | 1,987 | 0.74 |
| D5-S20 | 4h baseline window | 24.8 | −6.8 | 0 | 905 | 0.79 | 6.6 | −1.6 | 0 | 1,534 | 0.80 |
| D5-S21 | 6h baseline window | 26.4 | −5.2 | 0 | 691 | 0.84 | 6.9 | −1.3 | 0 | 1,226 | 0.84 |
| D5-S22 | 12h baseline window | 29.2 | −2.3 | 0 | 308 | 0.93 | 7.6 | −0.6 | 0 | 586 | 0.92 |

**D1 — Parameter sets.** Each repository parameter set was run on the common base cohort with all other settings at the reference, restricted to the clinician-validated union (Table 11). MIMIC-Code and trajPLT use every reference parameter and labelled exactly the reference stays; sepsis3-mimic and sepsis-3-labels missed four of them. The AI Clinician set lowered the MIMIC-III rate by 4.8 pp and the AI-enrichment set the eICU-CRD rate by 1.1 pp; both labelled almost only reference stays, losing 649 and 1,029 of them and gaining three each. Subscore shifts are shown in Figure 2.

**Table 11.** D1 parameter sets: detection rate of each repository parameter set on the common base cohort, difference from the reference (Δ, percentage points), septic stays of the reference that the set does not label (Lost) and septic stays it labels in addition (Gained), and Jaccard index $J$ of the septic stays against the reference (MIMIC-III 31.5 %, eICU-CRD 8.2 %).

| ID | Parameter set | Rate (%) | Δ | Lost | Gained | $J$ |
|---|---|---|---|---|---|---|
| *MIMIC-III* | | | | | | |
| D1-S1 | sepsis3-mimic [2] | 31.5 | 0.0 | 4 | 0 | 1.00 |
| D1-S2 | MIMIC-Code [3] | 31.5 | 0.0 | 0 | 0 | 1.00 |
| D1-S3 | sepsis-3-labels [4] | 31.5 | 0.0 | 4 | 0 | 1.00 |
| D1-S4 | AI Clinician [5] | 26.7 | −4.8 | 649 | 3 | 0.85 |
| *eICU-CRD* | | | | | | |
| D1-S5 | AI-enrichment [6] | 7.1 | −1.1 | 1,029 | 3 | 0.87 |
| D1-S6 | trajPLT [7] | 8.2 | 0.0 | 0 | 0 | 1.00 |

Figures 2–4 show the organ-level results. Figure 2 shows how the six repository parameter sets shift the individual SOFA subscores, as parameter sets alone (Tier 1) and as complete reconstructions (Tier 2). Figure 3 shows the same for the variants of D2–D5, and Figure 4 shows what share of each SOFA input is filled by imputation in the domains in which this share varies (D1, D2 and D4).

In the subscore figures (Figures 2 and 3), each cell gives the mean difference of an organ subscore or of the total SOFA from the reference in SOFA points, with blue for lower and red for higher values and a separate colour scale per panel. For D1, the reference is the reference configuration. For D2–D5, it is the reference variant of the domain, which is named in the panel title and not shown as a column. In Tier 1, each D1 parameter set runs on the fixed cohort, so that the difference reflects the parameters alone, whereas the Tier 2 reconstructions keep the cohort and windows of their repository, which also enter the difference. The eICU-CRD Tier 2 subscores do not depend on how infection is identified, so one column per repository is shown. The imputation figure (Figure 4) gives the fraction of SOFA inputs filled at each level (L1 derivation, L2 forward filling or interpolation, L3 constant default), and a blank cell marks a level that the variant disables. Aggregation and scoring act after imputation, so D3 and D5 would repeat the reference and are not shown. In all three figures, MIMIC-III panels are framed in blue and eICU-CRD panels in orange, and columns carry the identifiers of Section S2.1. The baseline and onset variants (D5-S13 to D5-S22) are omitted from the subscore figure because they change the label but not the subscores.

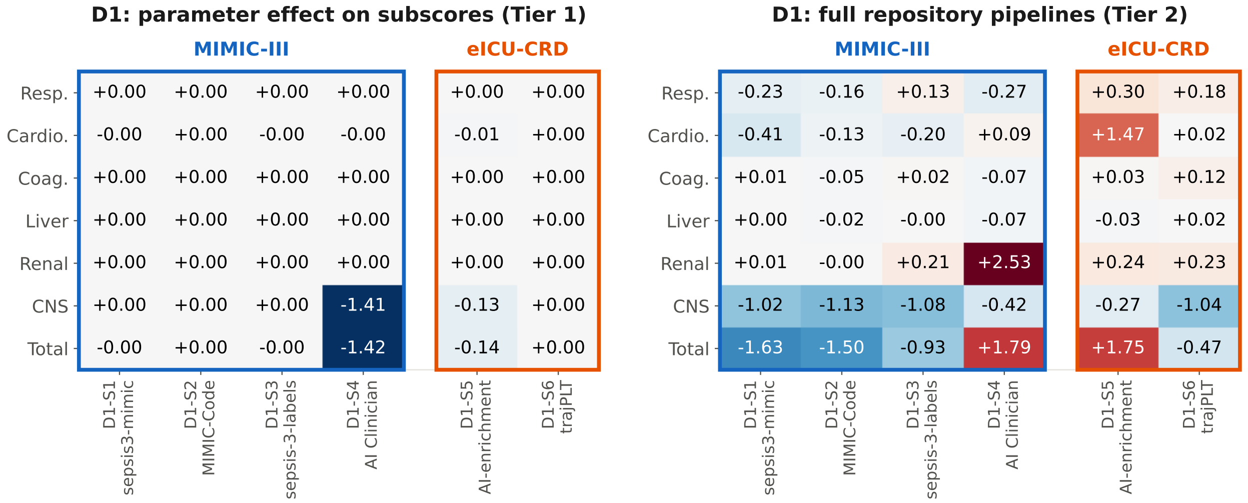


**Figure 2.** SOFA subscore differences of the six repository parameter sets (D1), in Tier 1 (left, each parameter set on the fixed cohort) and in Tier 2 (right, complete reconstruction of the repository pipeline). Reading conventions as described in the text.

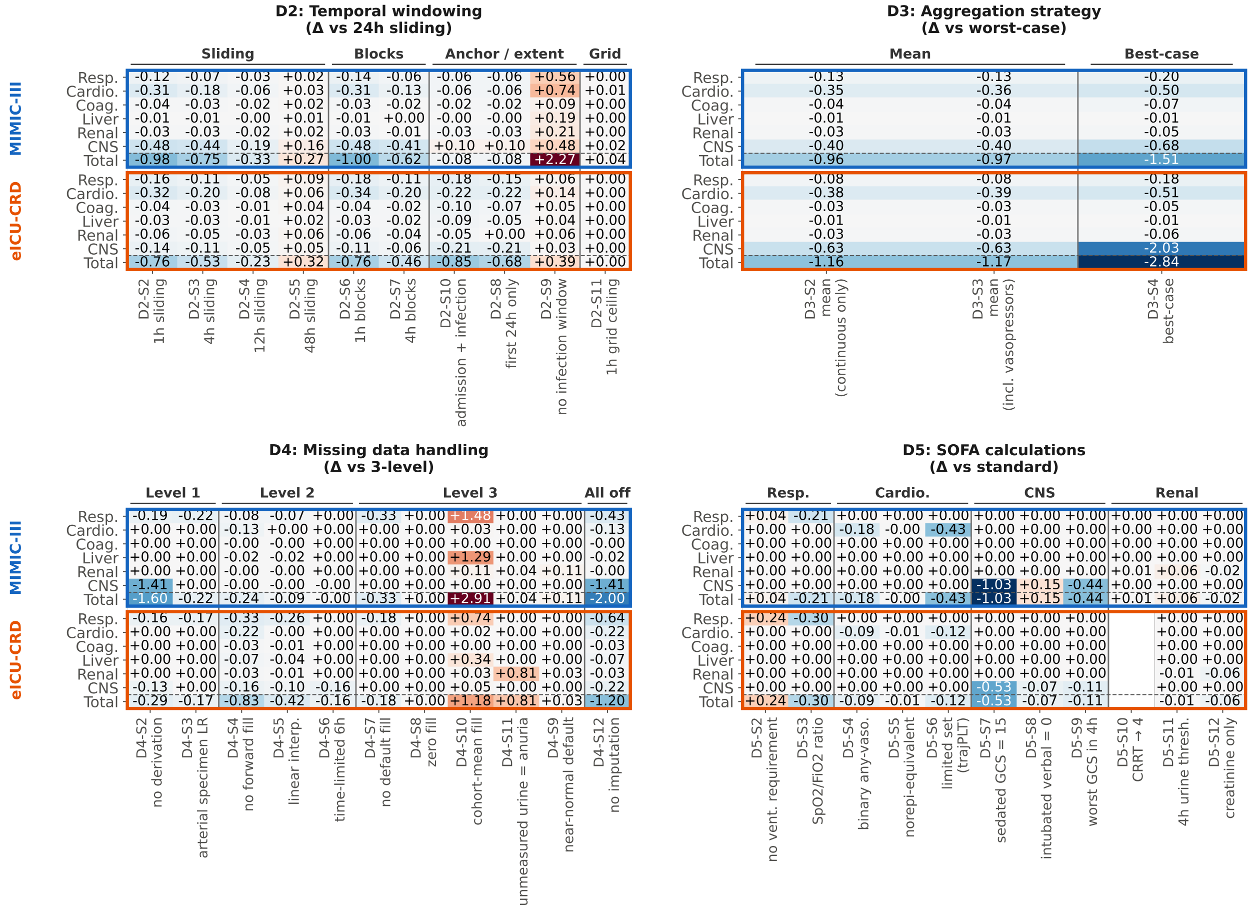


**Figure 3.** SOFA subscore differences of the variants of domains D2–D5 from the reference variant of each domain, with one panel per domain and MIMIC-III above eICU-CRD. Reading conventions as described in the text.

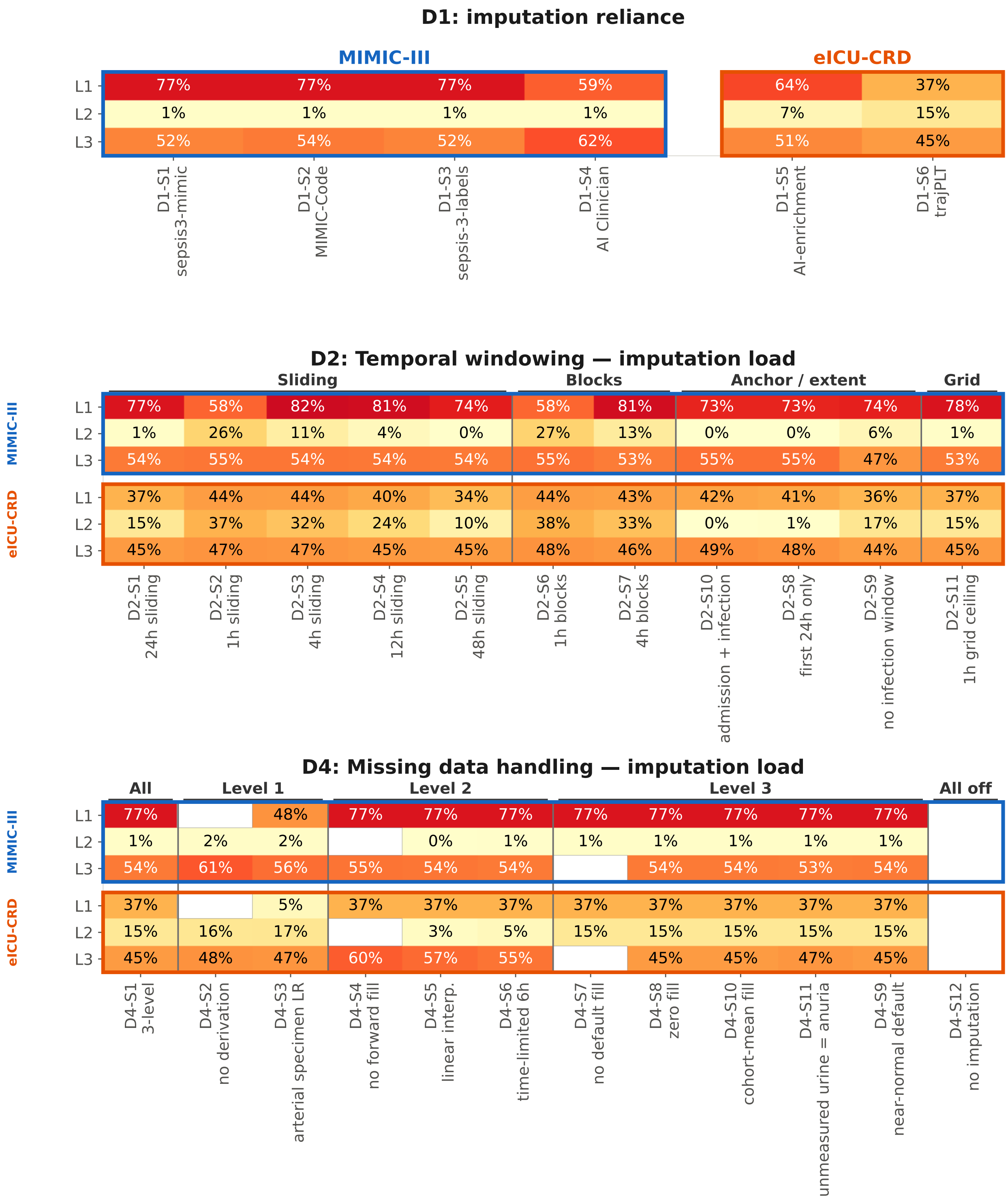


**Figure 4.** Share of SOFA inputs filled at each imputation level for the repository parameter sets (D1, top) and for the variants of temporal windows (D2) and missing values (D4), with MIMIC-III above eICU-CRD. Reading conventions as described in the text.

Exact detection rates for every Tier 1 and Tier 2 configuration are shown in Figure 1, Table 10 and the Tier 2 table of the main text and will be released in full with the code upon publication.

**Tier 3.** Four variants, one per processing domain, were combined in all six pairs per database. These are 1 h non-overlapping blocks (D2-S6), mean aggregation of continuous parameters (D3-S2), no derivation of missing values (D4-S2) and a zero SOFA baseline (D5-S15). Table 12 lists the pairwise combinations behind the interaction figure of the main text. The interaction is the observed deviation of the combination from the reference minus the sum of the two single effects, and values near zero indicate additivity.

**Table 12.** Tier 3 pairwise combinations: single-variant effects ($\Delta_A$, $\Delta_B$), their sum, the observed effect of the combination, and the interaction (observed − sum), in percentage points relative to the reference (MIMIC-III 31.5 %, eICU-CRD 8.2 %). Sums and interactions were computed from the unrounded rates.

| DB | Combination | $\Delta_A$ | $\Delta_B$ | Sum | Observed | Interaction |
|---|---|---|---|---|---|---|
| MIMIC-III | D2-S6 × D3-S2 | +12.6 | −10.1 | +2.5 | +12.2 | +9.8 |
| | D2-S6 × D4-S2 | +12.6 | −10.8 | +1.8 | −10.0 | −11.8 |
| | D2-S6 × D5-S15 | +12.6 | +27.1 | +39.6 | +28.0 | −11.6 |
| | D3-S2 × D4-S2 | −10.1 | −10.8 | −20.9 | −16.5 | +4.4 |
| | D3-S2 × D5-S15 | −10.1 | +27.1 | +17.0 | +23.4 | +6.4 |
| | D4-S2 × D5-S15 | −10.8 | +27.1 | +16.3 | +17.2 | +0.9 |
| eICU-CRD | D2-S6 × D3-S2 | +4.8 | −0.8 | +4.0 | +4.7 | +0.6 |
| | D2-S6 × D4-S2 | +4.8 | −2.3 | +2.5 | −2.3 | −4.9 |
| | D2-S6 × D5-S15 | +4.8 | +9.0 | +13.8 | +9.0 | −4.8 |
| | D3-S2 × D4-S2 | −0.8 | −2.3 | −3.1 | −2.8 | +0.3 |
| | D3-S2 × D5-S15 | −0.8 | +9.0 | +8.2 | +8.8 | +0.5 |
| | D4-S2 × D5-S15 | −2.3 | +9.0 | +6.7 | +8.6 | +1.9 |

### S2.4 Information Time, Time Origin, Documentation System and Infection Rule

All reviewed repositories and our reference configuration compute Sepsis-3 labels from measurement time (charttime), as if every value were available the instant it is measured. A real-time decision-support system sees a value only once it has been documented (storetime). Labels computed from measurement time therefore use values before such a system could have seen them and can overestimate how early sepsis is detectable. Our OMOP extension records both timestamps (`measurement_storetime`, `drug_exposure_storetime`; Supplement S1, Section S1.2). In the documentation-time setting, each value is placed in the SOFA time series at its measurement time but enters an aggregation window only once it is documented (`storetime` ≤ window end). A value measured inside a window but documented afterwards is withheld from that window.

Table 13 contrasts this documentation-time setting with the reference. In MIMIC-III the detection rate falls from 31.5 % to 26.5 % (−5.0 pp). Of the 4,223 septic stays of the reference, 918 (21.7 %) lose the label and 246 other stays gain it, leaving 3,551 septic stays, of which 3,305 are septic under both settings (Jaccard 0.74). In these 3,305 stays, the hour at which the label is first met is unchanged in 1,726 (52 %), later in 1,481 (45 %) and earlier in 98 (3 %). Where it is later, the median delay is 3 h, and it is at least 6 h in 415 stays. Results from the laboratory and microbiology tables carry no storetime in MIMIC-III and are treated as available at chart time, so this is a lower bound. The Bronze profile of MIMIC-III records the mean delay between measurement and documentation for every source parameter (column `avg_storetime_diff_secs`). Weighted by the number of observations, documentation followed measurement on average by 14 min for $PaO_2$, 18 min for ventilation, 28 min for $SpO_2$ and mean arterial pressure, 15–29 min for vasopressor rates with the exception of vasopressin (about 5.5 h), 30 min for urine output, about 40 min for GCS components, and 55, 81 and 129 min for platelets, creatinine and bilirubin charted at the bedside. eICU-CRD stores only minute offsets without a separate documentation time, so both settings coincide there by construction.

The same table reports three further one-setting contrasts. Anchoring the time origin $t_0$ at the first (any or SOFA-relevant) measurement instead of ICU admission changes rates by at most 0.9 pp in MIMIC-III and 2.2 pp in eICU-CRD. Requiring a four-day antibiotic course for suspected infection (D6-S2, Section S2.1) lowers the MIMIC-III rate from 31.5 % to 19.1 % ($-12.4$ pp). Including CareVue stays (cohort criterion C4) lowers the MIMIC-III rate to 26.2 % (CareVue alone: 22.4 %) under the culture-based infection definition, consistent with the underrepresented CareVue microbiology that motivates the MetaVision restriction (Section S2.1, D6).

**Table 13.** Documentation system, time origin, information time and infection rule. Sepsis-3 detection rate (number of ICU stays) and difference to the reference configuration in percentage points; each block varies one setting of the reference (MetaVision-only cohort, $t_0$ = ICU admission, measurement-time information, antibiotic + culture infection rule). eICU-CRD has no CareVue/MetaVision split and no separate documentation timestamp (its documentation time equals measurement time), and its reference reads infection from diagnosis codes, hence the empty and unchanged cells. The combined CareVue + MetaVision cohort also contains 74 stays documented in both systems, which belong to neither of the two subgroups.

| | MIMIC-III | | eICU-CRD | |
|---|---|---|---|---|
| **Setting** | **Rate (n)** | **Δ pp** | **Rate (n)** | **Δ pp** |
| *Documentation system (C4)* | | | | |
| MetaVision only* | 31.5 % (13,388) | ref. | — | — |
| CareVue only | 22.4 % (18,931) | −9.2 | — | — |
| CareVue + MetaVision | 26.2 % (32,393) | −5.3 | — | — |
| *Time origin $t_0$* | | | | |
| ICU admission* | 31.5 % (13,388) | ref. | 8.2 % (94,782) | ref. |
| First measurement (any) | 30.8 % (13,388) | −0.7 | 10.4 % (94,783) | +2.2 |
| First SOFA-relevant measurement | 30.7 % (13,388) | −0.9 | 10.4 % (94,783) | +2.2 |
| *Information time* | | | | |
| Measurement time* | 31.5 % (13,388) | ref. | — | — |
| Documentation time | 26.5 % (13,388) | −5.0 | — | — |
| *Infection rule* | | | | |
| Antibiotic + culture* | 31.5 % (13,388) | ref. | — | — |
| 4-day antibiotic course | 19.1 % (13,388) | −12.4 | — | — |
